# Severe Prenatal Shocks and Adolescent Health: Evidence from the Dutch Hunger Winter

**DOI:** 10.1101/2021.10.13.21264971

**Authors:** Gabriella Conti, Stavros Poupakis, Peter Ekamper, Govert E. Bijwaard, L.H. Lumey

## Abstract

This paper investigates impacts, mechanisms and selection effects of prenatal exposure to multiple shocks, by exploiting the unique natural experiment of the Dutch Hunger Winter. At the end of World War II, a famine occurred abruptly in the Western Netherlands (November 1944 - May 1945), pushing the previously and subsequently well-nourished Dutch population to the brink of starvation. We link high-quality military recruits data with objective health measurements for the cohorts born in the years surrounding WWII with newly digitised historical records on calories and nutrient composition of the war rations, daily temperature, and warfare deaths. Using difference-in-differences and triple differences research designs, we show that the cohorts exposed to the Dutch Hunger Winter since early gestation have a higher Body Mass Index and an increased probability of being overweight at age 18, and that this effect is partly accounted for by warfare exposure and a reduction in energy-adjusted protein intake. Moreover, we account for selective mortality using a copula-based approach and newly-digitised data on survival rates, and find evidence of both selection and scarring effects. These results emphasise the complexity of the mechanisms at play in studying the consequences of early conditions.

## 1 Introduction

The study of the long-term health effects of early life conditions has been at the core of life course epidemiology since its origins in the 1990s (Ben-Shlomo and Kuh, 2002). In the last decade, the economic literature has substantially contributed to the field, going beyond associational evidence to show causal impacts of early circumstances not only on health, but also on a wide variety of socioeconomic outcomes (see the reviews of Currie and Almond (2011), Almond et al. (2018), Prinz et al. (2018) and Conti et al. (2019)). While the field has been burgeoning, knowledge of the mechanisms through which early conditions affect life course outcomes is still scarce. Among the shocks most widely studied, prenatal malnutrition has often been proxied by exposure to a famine, caused by a war or other circumstances that affect entire cohorts in specific regions for limited periods of time.^1^ One of the most studied is the Dutch famine (also known as the “Hunger Winter”), which occurred at the end of World War II (November 1944 - May 1945) and was brief, unanticipated, and temporally and regionally defined.^2^

While previous work has convincingly shown lifelong health and socio-economic impacts of prenatal exposure to the Dutch Hunger Winter, starting with the influential paper by Ravelli et al. (1976), it has not been able to tackle some key issues, mostly because of data limitations. First, existing studies have mostly used a difference-in-differences methodology, comparing cohorts born in different periods in regions with varying exposure to the famine; hence, the relative importance of the multiple, co-occurring shocks which affected these cohorts – low-caloric and imbalanced nutritional content of the rations, but also harsh weather conditions and incessant warfare – has not been ascertained. As Van den Berg and Lindeboom (2018) note ‘If this point is ignored, this affects the interpretation of the famine as an effect through undernutrition’. Second, previous work has studied the impacts of the famine on the survivors, without systematically investigating the role played by selection effects. These limitations are shared by other studies on the long-term impacts of early shocks.

In this paper, we re-examine the original military recruits data used in the first, influential papers on the Dutch Hunger Winter (Ravelli et al. (1976) and Stein et al. (1972)), and we combine them with newly collected and digitised data from historical sources to provide a more comprehensive analysis of mid-term impacts, mechanisms and selection effects of a prenatal shock. The outcomes we study include: height and weight, which we use to compute indicators of underweight and overweight/obesity; chest circumference, which we use the construct a measure of body build (chest-height ratio); and intellectual disability. The novel data we have collected for this paper include: original information from the war rations on calories and nutrient composition of the diet; average daily temperature and number of deaths due to warfare; city-level demographic and socioeconomic characteristics; number of stillbirths, neonatal and post-neonatal deaths, and mortality until age 18.

Using these data, we improve upon previous famine studies in multiple ways. First, we use newly digitised data on the pre-war years to carefully select the cities in the control group, on the basis of common trends along a variety of demographic and socioeconomic indicators, rather than solely on the basis of geographical location. Second, we account for selective fertility, by focusing on the sample of women who were already pregnant at the start of the famine. In addition to our baseline difference-in-differences specification, we perform several robustness and placebo tests, including a triple difference design. Third, we use the newly collected information on caloric content and nutrient composition of the rations, average daily temperature, and number of deaths due to warfare, to disentangle the different mechanisms through which being in utero during the 1944-45 winter in West Netherlands affected health.^3^ Fourth, we account for selective survival, combining new information on the number of stillbirths, neonatal and post-neonatal deaths, and mortality until 18 years old (derived from historical records), with historical data on local occupational structure and availability of health care resources, within a flexible copula selection model. Throughout, we take good care of the inference by accounting for the moderate number of cities and for multiple hypothesis testing.

Our findings confirm that being exposed to the famine since the prenatal period is more harmful than being exposed only postnatally; most of the negative health impacts are concentrated among those exposed since early gestation.^4^ Affected cohorts have, on average, a significantly higher weight and a larger chest circumference, but not a greater height: hence, they have a higher prevalence of obesity. The choice of the control group matters: we show that the results obtained based on a control group of cities more broadly defined fail a simple placebo test. We then show that the obesity results are primarily driven by a change in the nutritional content of the rations – a reduction in the proportion of proteins – rather than by a dramatic fall in calories consumed, coupled with the stress deriving from exposure to warfare. Lastly, we confirm that conditioning on the sample of survivors leads to a downward bias of the estimated famine effects, and provide evidence of two-sided selection and scarring: on the one hand, those underweight and overweight are both less likely to survive than those with a normal weight; on the other hand, even after accounting for selection, the famine caused significant scarring, with a 11.67% and a 35.71% higher prevalence of overweight and obesity, respectively, among the survivors. Additionally, once we account for selection we are able to detect a significant impact of the famine among those exposed since middle gestation on the likelihood of having an intellectual disability by 46.67%, which can be rationalized by the fact that those more likely to be mentally impaired were less likely to survive.

Our work provides several contributions to the literature on the early origins of health. First of all, it revisits and significantly expands - by using newly digitised data extracted from historical World War II documents and modern econometric techniques - some of the most influential epidemiological papers in the field of the developmental origins of health. Second, it provides a template for addressing issues usually left unaccounted for by previous studies, such as selective fertility, survivor bias and mechanisms. Third, it belongs to the small group of studies which elucidate the importance of the “missing middle”, i.e. the period between the early shock and the adult outcomes which is also less usually studied, for the lack of suitable data.

The rest of the paper is organised as follows. Section 2 presents the Dutch famine of 1944-1945 and discusses the relevant literature on the relationship between *in utero* malnutrition and adult outcomes. Section 3 describes the data sources and Section 4 describes the econometric specifications used in this study. Section 5 presents the results and Section 6 concludes.

## 2 Background

### 2.1 The Dutch Hunger Winter of 1944-1945

Towards the end of the 1940-1945 Nazi occupation of the Netherlands during the World War II, food – especially in the big cities – was distributed with rations (which included bread, potatoes, meat, butter and other fats). During the winter of 1944-1945, the still occupied part of Netherlands experienced a severe famine as a result of the Nazi blockade, triggered by the Dutch national railways strike to facilitate the Allied liberation efforts. The situation became even worse due to the low temperatures in the winter period, the freezing of the canals, and the military stalemate of the Allied forces on the Dutch front. While throughout the occupation the food rations were maintained at around 1,800 calories per day per person, they dropped to below 1,000 calories per day by November 1944 and to 500 calories per day by April 1945 (consisting mainly of bread and potatoes). The famine ended with the liberation of the occupied part in early May 1945.

This extreme shortage was experienced mainly in the Western Netherlands; in the North and East the rationing of the food was far more limited (see Lumey and van Poppel (1994) for further details), while the South was mostly already liberated. With 3.5 million people (of a total population of 9.3 million) living in the cities of the West – the most affected by the famine – the effects of this shortage were particularly severe. The estimated war-related excess deaths vary between 15,000 and 25,000 (see Ekamper et al. (2017) for a discussion of various estimates). While the famine affected the entire population living in the Western Netherlands, more than 40,000 individuals were exposed *in utero*, making it a suitable “natural” experiment to study the consequences of prenatal shocks.

### 2.2 In Utero Shocks and Adult Health Outcomes

The World War II had devastating consequences for the civilian populations across Europe, as found in many studies using retrospective data on childhood exposures.^5^ War can affect physical and mental health via multiple channels, such as experience of hunger, dispossession, absence of the father, and stress from combats (Kesternich et al., 2014). Examining self-reported hunger episodes in the Netherlands, Germany and Greece, Van den Berg et al. (2016) find significant effects on height for men (but not for women), with the reduced form effect being a 0.7 cm reduction, and the instrumental variable estimate (using the propensity to report hunger) a, rather substantial, 3.4 cm decline.

In the context of World War II, many studies have examined the Dutch famine of 1944-1945. The landmark study of Ravelli et al. (1976) used data on military recruits examined at 18 years old, and found an effect for those with prenatal exposure on the likelihood of obesity (defined as the weight to height ratio being equal to or greater than 120 percent). More specifically, the authors found that those exposed in the first half of pregnancy had higher obesity rates, while those exposed in the last trimester of pregnancy and first months of life had lower obesity rates. Using the same data on military recruits, Stein et al. (1972) failed to find any significant association between in utero malnutrition and mental performance at age 18. In other two cohort studies of men and women followed from birth to late middle age, prenatal famine exposure was associated with increased BMI and waist circumference in women (Ravelli et al., 1999; Stein et al., 2007). Two further studies evaluating the effects on cognition, instead, found contradictory results.^6^ Two recent papers in economics have also studied the effects of the Dutch famine. Scholte et al. (2015) use high-quality register data and find higher hospitalisation rates in the years before retirement if exposure occurred in middle or late gestation, and a significant decrease in the likelihood of being employed at age 55 for those exposed in early gestation, which they interpret as a proxy for decline in cognitive ability. Portrait et al. (2017) use the Longitudinal Ageing Study Amsterdam (which includes cohorts born in 1930–1945), and find a 4 cm significant reduction in height for both males and females exposed between gestation and age 2.

Other papers have studied episodes of famine experienced in other countries at the time of WWII. Jürges (2013) finds negative effects of the German famine (proxied by birth cohort-month exposure) on both education and labour market outcomes, stronger for early pregnancy than for late pregnancy exposure. Also for Germany, Kesternich et al. (2015) find higher food expenditures among lower-income adults who experienced hunger in childhood (self-reported episodes validated with comparisons with to office food rations) – suggesting a possible behavioural pathway through which the health effects could manifest. Akbulut-Yuksel (2017), instead, studies the effect of the intensity of WWII destruction in Germany and finds that individuals who were exposed during the prenatal and early postnatal period have higher BMI and are more likely to be obese as adults – an effect that she attributes to early life malnutrition. Neelsen and Stratmann (2011) examine the effects of the Greek famine 1941-1942 using census data and find negative impacts on education and literacy for those exposed in infancy; additionally, they find stronger effects for the urban-born cohorts compared to the rural-born ones, suggesting, like other studies, that the famine was mostly experienced in the urban centres.^7^ Atella et al. (2020) exploit Nazi raids on municipalities in Italy during WWII and find that workers exposed to raids in utero have worse labor market outcomes. Allais et al. (2021) show that both the intensity of WWII exposure in France (measured as the number of military casualties) and self-reported episodes of hunger are associated with worse health outcomes for a sample of women over 20s.

Hence, while multiple studies have examined long-term impacts of exposure to famine, one characteristic common to all of them is the analysis of sample of survivors. In this case, the exposed cohort is subject to two effects working in opposite directions: on the one hand, those who survive are ‘scarred’ and thus have worse health; on the other hand, those at the bottom of the health distribution are less likely to survive, resulting in left truncation and a population with better health. Bozzoli et al. (2009) argue that an environment with high infant mortality favours the selection effects to dominate, whereas in settings with better conditions and lower mortality scarring is more evident, i.e. the survivors are generally shorter and less healthy. However, most studies have not attempted to account for selection; among the few exceptions there are Gørgens et al. (2012) and Lindeboom et al. (2010), who use the Historical Sample of the Netherlands with continuous life histories and find that most of the selection effects take place at early ages, and that their impacts can be substantial.

## 3 Data and Sample Definition

### 3.1 Military Recruits Data

The primary data source for our analysis is the original military recruits data used in the well-known studies of Stein et al. (1972) and Ravelli et al. (1976) in the 1970s. This includes all men born in the Netherlands in the period 1944-1947 who were examined in the military centers at age 18. In addition to the results of the medical examinations, the data contains the exact date and place of birth, and basic demographic information (father’s occupational status and family size).

As mentioned in section 1, to account for selective fertility, we only include in our analytical sample those individuals who were already conceived at the start of the famine, i.e. those born up to July 1945. We then define three treatment groups: early exposure starting in the first trimester (born May-July 1945), middle exposure starting in the second trimester (born February-April 1945), and late exposure starting in the third trimester (born November 1944-January 1945). The control group includes those exposed only postnatally, in the first months of life (born May-October 1944).^8^ Figure 1 shows the time periods corresponding to each of the three treatment groups and the control group; the famine period is indicated by the red vertical lines. It is worth noting that, given the 6-month duration of the famine, nobody was exposed for the full duration of the pregnancy: those exposed since mid- and late-gestation had also some postnatal exposure, and those exposed since the first trimester were born after liberation and not exposed in the third trimester. Defining exposure by counting backwards from the date of birth, as opposed to counting forwards from the date of conception, has well-known problems (Currie and Rossin-Slater, 2013). However, we do not consider this is an issue in our application, given that the famine did not affect gestation length (Stein and Susser, 1975), as we confirm in our birth sample, which is based on high-quality hospital records (column (7) of Table 7).

**Figure 1:**
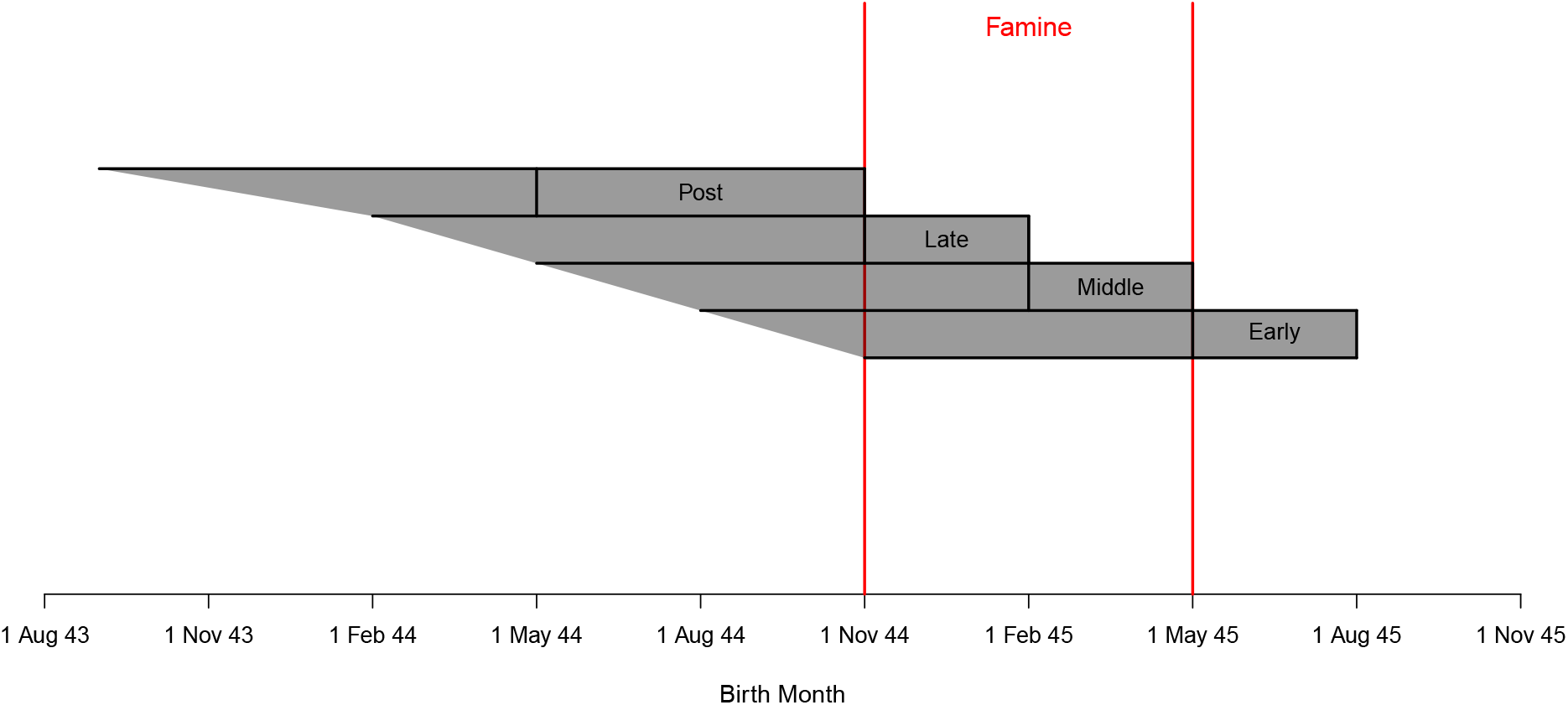
Treatment and control groups definition Note: The figure shows the time periods corresponding to each of the three treatment groups and the control group, with the birth months for each group enclosed in the respective grey boxes labelled “Post” (May-October 1944), “Late” (November 1944-January 1945), “Middle” (February-April 1945), “Early” (May-July 1945). The pregnancy period corresponding to each birth month is shaded in grey. The red vertical lines enclose the famine period.

Additionally, as also mentioned in section 1, we take great care in selecting the cities included in the analytical sample–differently from what done in most of the literature. We follow the following steps. First, given that the famine historically affected more the cities (Banning, 1946; De Jong, 1981), we restrict our sample of interest to the 46 municipalities with a population greater than 25,000 inhabitants on January 1, 1940. Second, using data from the Historical Ecological Databank of the Netherlands (Boonstra, 2016), we exclude: (a) 4 municipalities classified as rural on the basis of their population dispersion pattern; (b) 13 municipalities where the population underwent major changes in size since 1930 (i.e. either increased by more than 50% over a decade or decreased after the onset of the war).^9^ Table 1 lists the excluded and retained cities, and their allocation into Famine or control areas. Third, we test and fail to reject that the remaining 29 cities follow the same trends before the start of WWII in several health and demographic outcomes (postnatal mortality rate, crude birth rate, crude death rate, crude marriage rate, infant mortality rate, mortality rate 1-14 years old, mortality rate 15-39 years old, mortality rate 40-59 years old, mortality rate 60+ years old, using data compiled from published monthly statistics by city from the Dutch Central Bureau of Statistics (CBS, 1935-1947). The results are presented in Figures B1 to B9 in the Appendix: here we also see that, in the case of the non-selected cities, we reject the null hypothesis that they were on the same trends for many outcomes.^10^ Hence, we work with a consistently defined group of 7 treatment (in the West) and 22 control cities (6 in the West, 7 in the North-East, and 9 in the South).

**Table 1:**
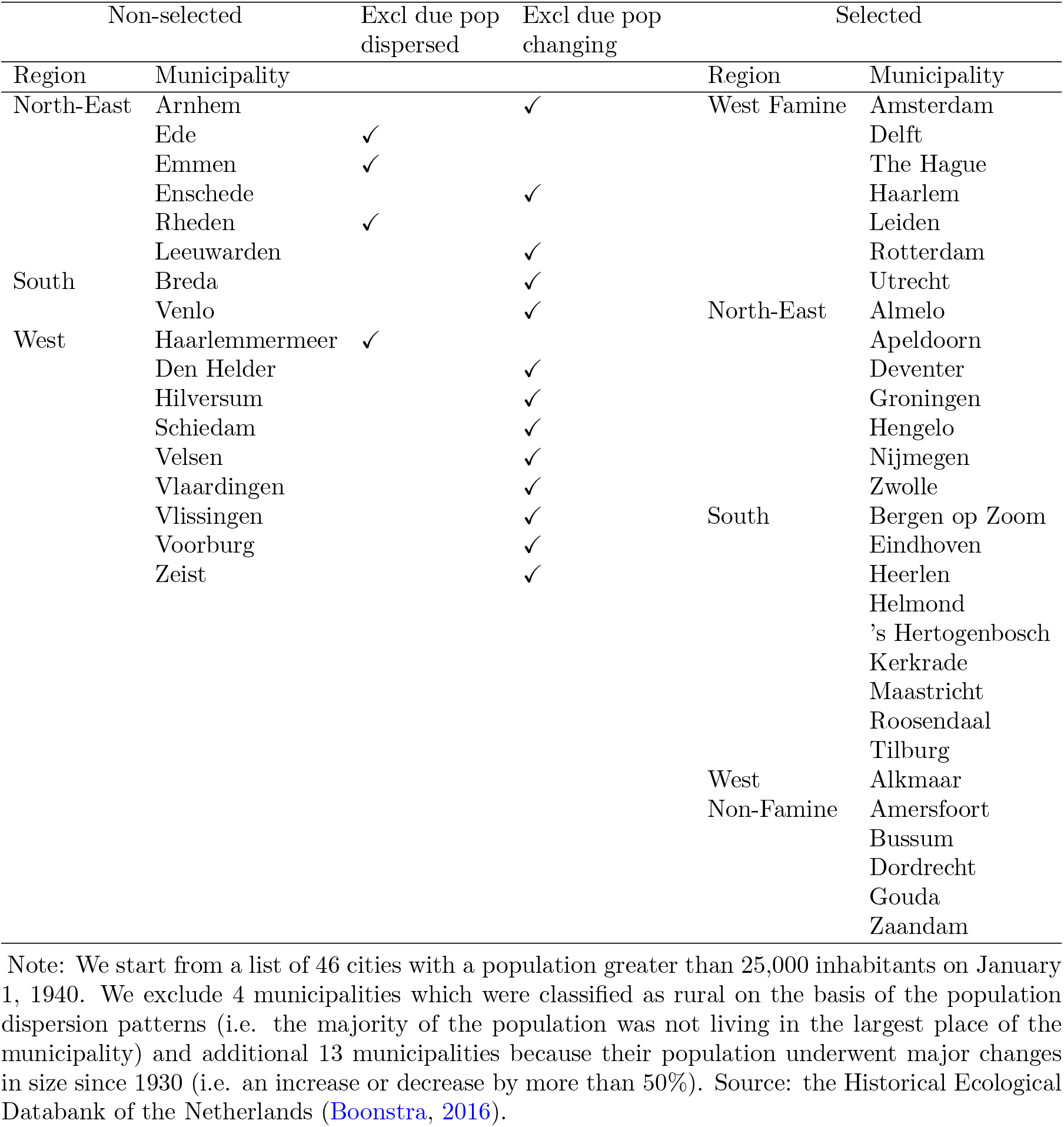
Selection of Cities Summary

### 3.2 Newly Digitised Historical Data

To investigate possible mechanisms, we enrich the military recruits data by merging them (using the date and city of birth) with newly digitalised information on caloric content and nutrients composition of the rations, temperature, civilian deaths from warfare and the dates in which the cities in the South were liberated by the Allied forces. First, we extract the data on caloric content and nutrients composition from the official war information on the rations (Departement van Landbouw en Visserij, 1946); this information is available at weekly level for the West and at monthly level for the North-East and South of the Netherlands. Figure 2 shows the distribution of calories and shares of protein, fat and carbohydrates for each trimester by month of birth, separately for the West, North-East and South.^11^ The Figure clearly shows that the drop in calories during the famine was accompanied by a drop in the protein share at the end of gestation among those with early exposure in the West, and that the liberated South had rations with higher caloric intake and greater nutritional value.

**Figure 2:**
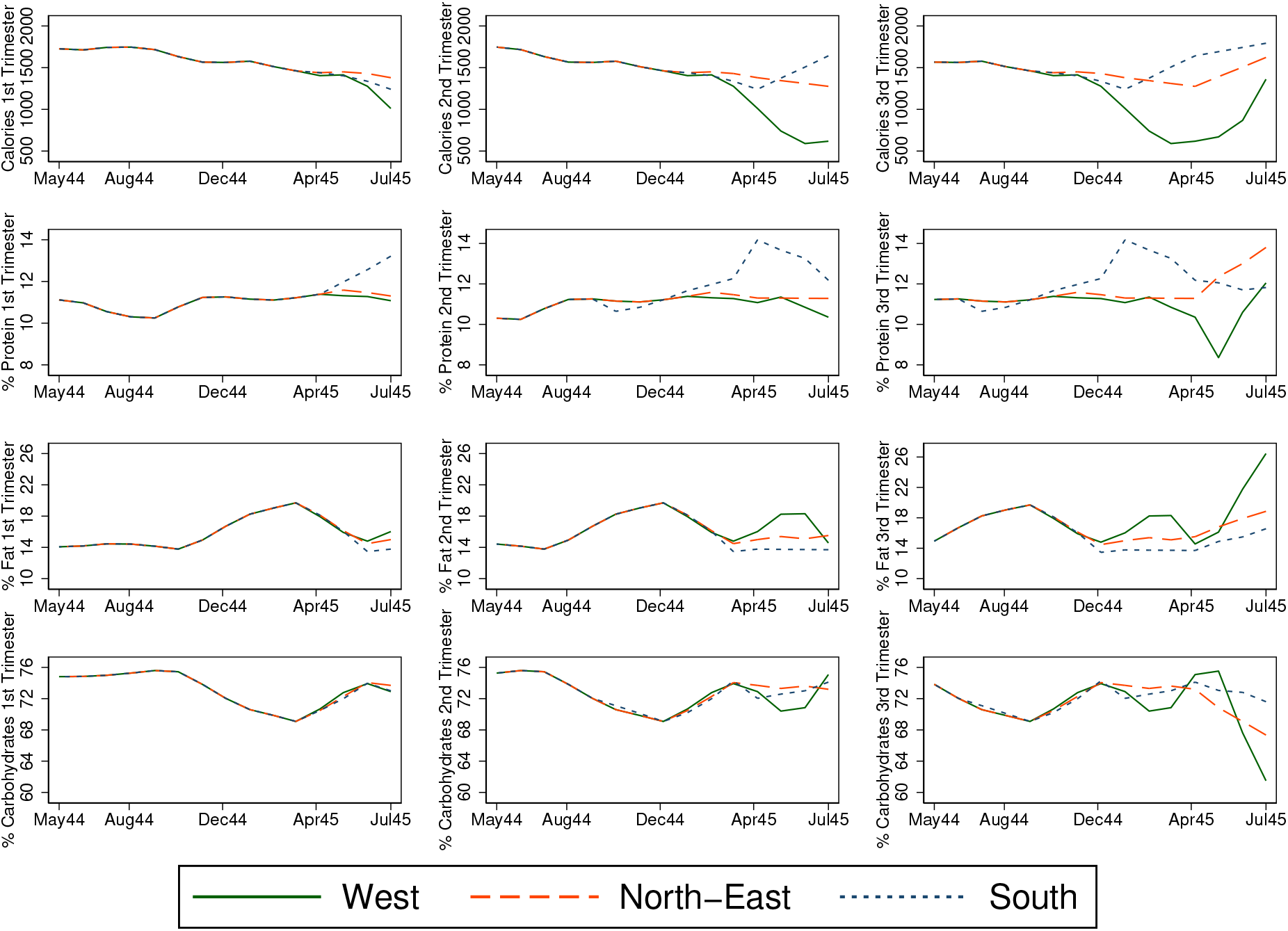
Rations for each trimester by month of birth and region Note: Data on calories (first row) and macronutrients shares (% protein, fat and carbohydrates in second, third and fourth row, respectively) at weekly level for the West and at monthly level for the North-East and South, by month of birth (shown on the horizontal axis). Source: official war information on the rations. Note that West here includes both the treated and the control cities. Shares are calculated as *P rotein share* = 4 *× P rotein*(*grams*)*/Calories*(*kcals*), *F at share* = 9 *× F at*(*grams*)*/Calories*(*kcals*) and *Carbohydrates share* = 100 *−* (*P rotein share* + *F at share*).

Second, we extract temperature information for the period of interest, from the archives of the Royal Netherlands Meteorological Institute (KNMI, 2018). At the time, only three meteorological stations were operating in the Netherlands, thus we use Inverse Distance Weighting (Pebesma, 2004) to interpolate the average daily temperatures for each city in our sample, separately for each month. For illustrative purposes, Figure D1 presents an example of the interpolated data for two months, December 1944 and May 1945, based on the data from the meteorological stations (the red squares). The heatmap shows the predicted temperatures across the Netherlands, as indicated in the legend, with each city (the black dots) receiving a value depending on its location. The famine occurred during a winter that, from a purely climatological perspective, was not unusually harsh overall except for a period at the end of January 1945,^12^ when parts of the country were still under occupation and some others (the South) were being liberated.

Third, we extract from the CBS registries information on the number of civilian deaths from war-related causes.^13^ Deaths due to warfare are all deaths classified with code “197 - Deaths of civilians due to operations of war” (within the main category “XVII – Violent or Accidental Deaths”) according to the International List of Causes of Death, Revision 5 (ICD-5) of 1938 (CBS, 1935-1947); this classification allows to better separate direct and indirect mortality from the war (Jewell et al., 2018). These civilian deaths were in part the result of bombings by the Allied Forces (Ekamper, 2020). During the occupation, the Allied Forces carried out around 600 bomb attacks on Dutch territory (Korthals Altes, 1984; NIOD, 2018), aimed at strategic targets, such as ports, bridges, and railways. Most bombings caused no or relatively few deaths among the civilian population; however, in few cases the attacks caused large numbers of civilian casualties (because of errors or missed targets, for example in Nijmegen there were nearly 800 victims on February 22nd, 1944, and in The Hague there were around 550 victims on March 3rd, 1945). Figure 3 shows the number of civilian deaths due to war operations for each birth month by city for the period May 1944 to July 1945; each plot has three lines, showing the civilian deaths for each trimester. The warfare affected cities in all three regions (West, North-East and South) within our study period for all three trimesters, allowing us to explore any trimester-specific effects.

**Figure 3:**
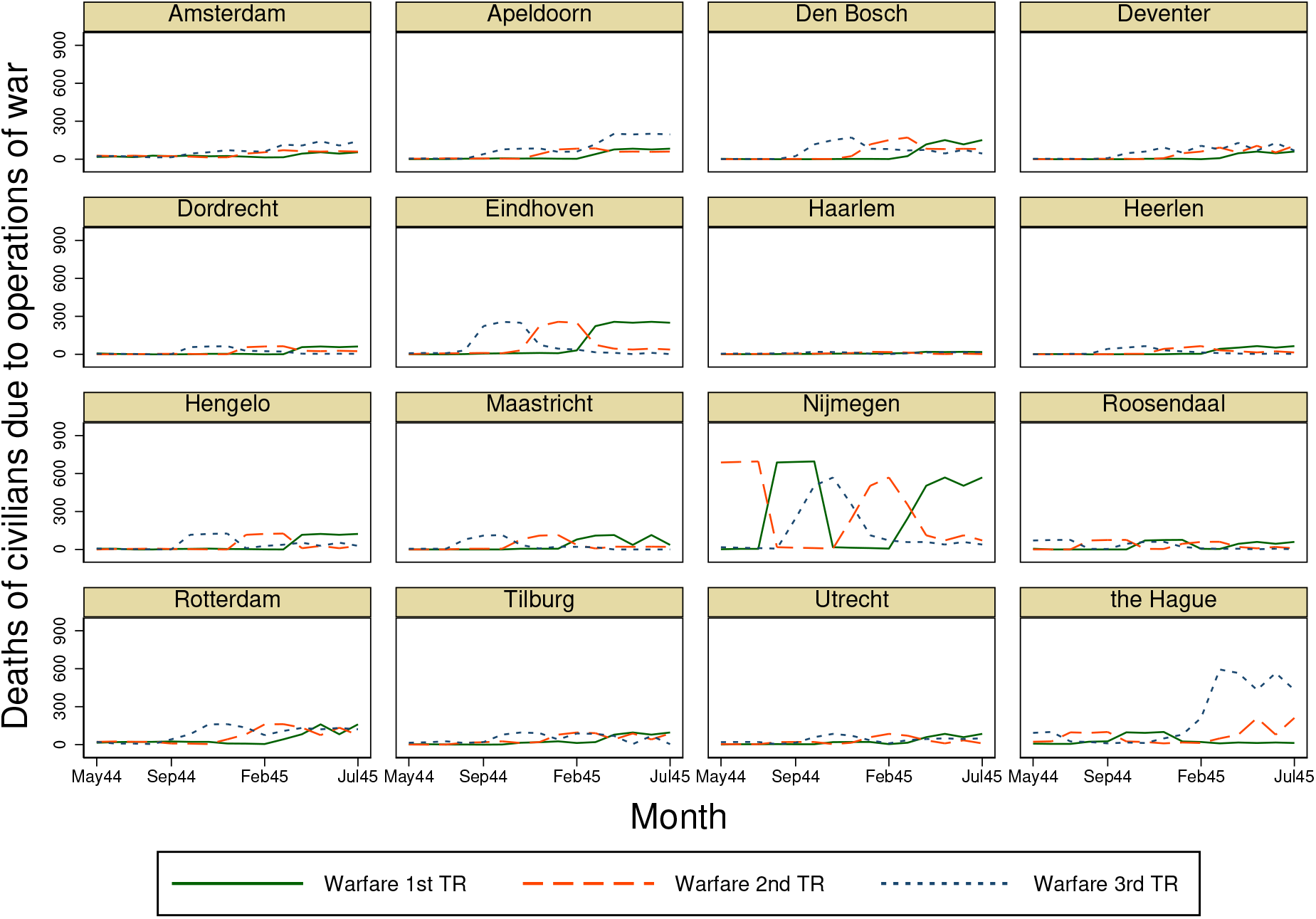
Civilian deaths due to war operations during pregnancy for each trimester by month of birth and city Note: Results for those born between May 1944 and July 1945 (month of birth displayed on the horizontal axis). The cities of Alkmaar, Almelo, Amersfoort, Bergen op Zoom, Bussum, Delft, Gouda, Groningen, Helmond, Kerkrade, Leiden, Zaandam, and Zwolle had little variation and are not shown here. Source: CBS registries, deaths with code “197 – Deaths of civilians due to operations of war”.

Fourth, we extract from historical sources information on the dates in which each city in the South was liberated from the Nazi by the Allied forces: 14 September 1944 (Maastricht), 17 September 1944 (Heerlen), 18 September 1944 (Eindhoven), 23 September 1944 (Helmond), 5 October 1944 (Kerkrade), 27 October 1944 (Den Bosch and Tilburg), 29 October 1944 (Breda), 30 October 1944 (Bergen and Roosendaal), 1 March 1945 (Venlo). We merge this information by date of birth (‘number of days since liberation’) to account for the fact that conditions in the South changed during the study period. Table 2 shows the descriptive statistics of these additional data merged with the recruits sample.^14^

**Table 2:** Newly Digitized Historical Data, Descriptive Statistics

| Variable | Level | Mean | Std. Dev. | Min | Max |
| --- | --- | --- | --- | --- | --- |
| Temperature (°C) 1st TR | Monthly | 8.54 | 4.75 | 1.70 | 16.70 |
| Temperature (°C) 2nd TR | Monthly | 8.07 | 4.91 | 1.70 | 16.70 |
| Temperature (°C) 3rd TR | Monthly | 9.88 | 4.70 | 1.70 | 16.70 |
| log(Warfare 1st TR + 1) | Monthly | 2.17 | 1.56 | 0.00 | 6.55 |
| log(Warfare 2nd TR + 1) | Monthly | 2.57 | 1.64 | 0.00 | 6.55 |
| log(Warfare 3rd TR + 1) | Monthly | 2.92 | 1.71 | 0.00 | 6.39 |
| Calories 1st TR (1,000s) | Weekly | 1.49 | 0.26 | 0.59 | 1.75 |
| Calories 2nd TR (1,000s) | Weekly | 1.34 | 0.35 | 0.59 | 1.75 |
| Calories 3rd TR (1,000s) | Weekly | 1.31 | 0.37 | 0.59 | 2.05 |
| Protein Share 1st TR | Weekly | 11.11 | 0.60 | 10.25 | 14.18 |
| Protein Share 2nd TR | Weekly | 11.07 | 0.82 | 8.66 | 14.18 |
| Protein Share 3rd TR | Weekly | 11.28 | 0.94 | 8.66 | 14.18 |
| South Weeks Liberated | Weekly | 1.70 | 8.57 | 26.00 | 44.43 |

Note: The statistics refer to the cohorts in the analytical sample (May1944-July1945 in the selected cities). The number of observations for all the variables is 40,950. The calories and protein shares are transformed into negative values for the estimation.

Fifth, we calculate survival rates up to age 18 (proportion of those born who are alive at age 18) by combining the information in the recruits data with data on live births and deaths (still births^15^, and deaths *<*6, 7-29, 30-89, 90-364 days and 1-18 years) by sex, region and month from Stein et al. (1975) (Table 1), which is based on extensive follow-up of pre-determined cohorts from birth to age 18.^16^ Lastly, we extract from the Historical Ecological Databank (HED) of the Netherlands (Boonstra, 2016) the following city-level pre-war information (measured in 1930), to help identification in the selection models: medical staff per 1,000 of population, the share of inhabitants of the largest place in the municipality (over the total number of inhabitants) and the number of religious groups with *≥*25 inhabitants in 1930. The choice of these exclusions has been guided by the following considerations. First, given that the Nazi embargo closed the imports also of medications (and of other vital supplies in addition to food) and that the disease environment also worsened (Van den Berg and Lindeboom, 2018; Banning, 1946), it is plausible that individuals born in cities with greater availability of medical infrastructures were more likely to survive. Second, mortality in small towns was lower (Banning, 1946). Third, it has been documented that religious associations (in particular the Inter-Church Council, I.K.O.) were very active in helping the vulnerable population at risk of starvation (Banning, 1946).

### 3.3 Birth Hospital Data

We complement the analysis of age 18 outcomes based on the military recruits data with unique birth data from the hospital records of five cities: Amsterdam, Rotterdam and Leiden (West), Groningen (North) and Heerlen (South) (Stein and Susser, 1975).^17^ Although at the time less than half of the births were taking place in hospitals, the admission procedures remained unchanged during the war (Stein and Susser, 1975). We use data on all births that occurred between May 1944 and July 1945. For each birth record, we have information on the sex of the newborn, weight, length, head circumference, along with placenta’s weight, gestational age, and mother’s age at birth.

## 4 Econometric Framework

We use a difference-in-differences design to estimate the impact of the famine on various health outcomes at age 18. Our main estimating equation is:

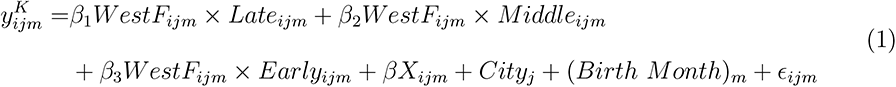

where 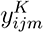 is one of the following outcomes *K* for person *i*, born in city *j* in month *m*: *Height*, *Weight*, *BMI* (*kg/m*^2^), *Overweight* (*BMI ≥* 25), *Obesity* (weight/height ratio *≥* 120% –we use the definition as in Ravelli et al. (1976) for comparison purposes), *Underweight* (*BMI <* 18.5), *Chest-Height Ratio* (a measure of body size used in Costa (2004)), *Intellectual Disability* (ICD-6 325, primary or secondary diagnosis, as defined in Stein et al. (1972)). *WestF_ijm_* is a binary indicator which takes value 1 for those born in the West Famine region. *Late_ijm_*, *Middle_ijm_* and *Early_ijm_* are binary indicators which take value 1 for those born between November 1944 and January 1945, between February 1945 and April 1945, and between May 1945 and July 1945, respectively (see Figure 1) – i.e. with exposure starting from the third, the second and the first trimester.

The reference group includes those born in the selected cities in Non-Famine West, North-East and South regions (see Table 1), between May and October 1944 – hence with only postnatal exposure to the famine. We do not include those conceived during or after the famine, in view of the significant reduction first, and subsequent increase, in conceptions and births, particularly among those with low socioeconomic status (Stein et al., 1975). Finally, *City_j_* and (*Birth Month*)*_m_* are fixed effects, and *X_ijm_* is a vector of controls (binary indicators for father’s occupational status, number of older brothers, birth order, and religion).^18^

The main parameters of interest are *β*_1_*, β*_2_ and *β*_3_, the interaction terms of *WestF_ijm_* with the three exposure dummies, which can be interpreted as an Intention-to-Treat (ITT) effect. In all estimations we use clustered standard errors;^19^ given the relatively small number of cities, we follow the recommendation in Cameron et al. (2008) and compute wild cluster bootstrap standard errors, reporting the corresponding p-values (Roodman et al., 2019). To account for the multiplicity of hypotheses tested, we use the Romano and Wolf (2005, 2016) procedure.^20^

After having estimated our baseline specification, we perform two additional analyses to allow for the fact that seasonal effects might differ between famine and non-famine regions. First, we estimate the same model as in Equation 1, but on a sample of cohorts born two years later (i.e. between May 1946 and July 1947), as a placebo regression. Second, we estimate the following triple difference (difference-in-difference-in-differences) specification:

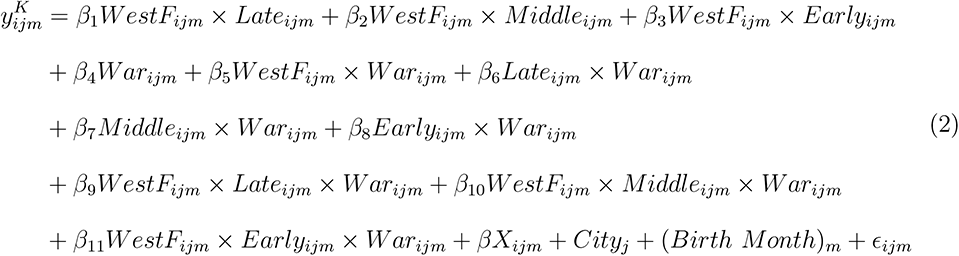

where the variables *WestF* and *Late*, *Middle* and *Early* are defined as before. The variable *War* is a dummy variable taking value 1 for those born during the war period (May 1944 - July 1945) and 0 for those born two years after (May 1946 - July 1947). The coefficients of interest in Equation 2 are those of the triple interactions: *β*_9_, *β*_10_ and *β*_11_.

As previously mentioned, we complement the analysis of the health outcomes at age 18 with that of the birth outcomes, using the data on the six hospitals. The main estimating equation is:

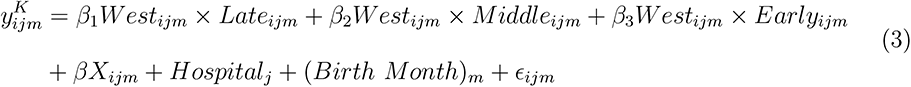

where 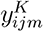 is one of the following birth outcomes for child *i* born in city *j* in month *m*: *Birth Weight*, *Low Birth Weight* (birth weight *<* 2, 500 grams), *Birth Length*, *Head Circumference*, *Placenta Weight*, *Gestation Age*, and *Sex Ratio*. The analytical sample includes only male births (for comparison with the main results), except for *Sex Ratio* which includes both males and females. *Late_ij_*, *Middle_ij_*, *Early_ij_* are defined as before. *West_ij_* is a dummy variable taking value 1 for those born in one of the hospitals in Amsterdam, Leiden and Rotterdam, and 0 for those born in Groningen and Heerlen. *Hospital_j_* and (*Birth Month*)*_m_* are the hospital and month of birth fixed effects and *X_ij_* includes mother’s age as control.

Finally, we assess the robustness of the results to accounting for selection into survival up to age 18.^21^ We use a selection model and depart from the bivariate normality assumption on the error terms by using copulas;^22^ in our implementation we choose among different copulas (Gaussian, FGM, Plackett, Clayton, AMH, Frank, Gumbel, and Joe) using the Bayesian Information Criterion (BIC). The estimating outcome equation is 1, and the selection equation is:

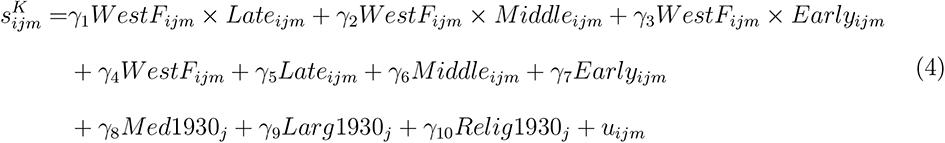

where 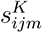 indicates whether the individual is alive at age 18 (constructed as explained in section 3.2), and the error terms *E_ijm_* and *u_ijm_* have a joint distribution based on one of the proposed copulas. For example, in the case of a Gaussian copula, the joint distribution is a bivariate Normal distribution, commonly used in many applications of sample selection models. The rest of the variables are defined as before, and *Med*1930, *Larg*1930 and *Relig*1930 are the 1930 city-level variables described in section 3.2.

## 5 Results

### 5.1 Main results

Our baseline results, based on the difference-in-differences specification (Equation 1), are presented in Table 3. All coefficients are from linear regression models, so that they can be interpreted directly as marginal effects. The individuals exposed since early gestation (i.e. trimesters 1 and 2 only, as the famine period was limited to 6 months) have a significantly higher weight (by 652 grams w.r.t a control mean of 67.6 kg), BMI (by 0.2 w.r.t a control mean of 21.5), obesity (using the Ravelli definition,^23^ by 0.6 p.p. w.r.t a control mean of 1.4%), and chest-height ratio (a 0.010 increase w.r.t a control mean of 0.492); they are also significantly less likely to be underweight (by 1.3 p.p. w.r.t a control mean of 5.6%). The individuals exposed since mid-gestation (i.e. trimesters 2 and 3) have only a significant increase in chest-height ratio (a 0.008 increase w.r.t a control mean of 0.492) and in intellectual disability (by 1 p.p. w.r.t. a control mean of 3%). The individuals exposed since late gestation (i.e. trimester 3 only) have significantly *lower* weight, overweight and obesity (using the Ravelli definition), as compared to those with exclusive postnatal exposure. All these effects are robust to controlling for multiple hypothesis testing by using the Romano and Wolf (2005) step-down method. We are unable to detect any impacts for any exposure group for height.

**Table 3:** Difference-in-Differences Estimates of the Dutch Hunger Winter Effects on Age 18 Outcomes

|  | (1) | (2) | (3) | (4) | (5) | (6) | (7) | (8) |
| --- | --- | --- | --- | --- | --- | --- | --- | --- |
|  | Height | Weight | BMI<br>(Weight/<br>Height<sup>2</sup>) | Overweight<br>(BMI<br>≥25) | Obese<br>(Weight/<br>Height>120%) | Underweight<br>(BMI<<br>18.5) | Chest/<br>Height<br>Ratio | Intellectual<br>Disability<br>(ICD-6 325) |
| WestF×Late | -0.099<br>(0.150) | -0.350*<br>(0.193) | -0.092<br>(0.058) | -0.010*<br>(0.005) | -0.006**<br>(0.003) | 0.001<br>(0.006) | 0.000<br>(0.001) | 0.002<br>(0.006) |
| WestF×Middle | -0.384<br>(0.305) | -0.334<br>(0.445) | -0.012<br>(0.106) | -0.006<br>(0.006) | 0.001<br>(0.004) | -0.003<br>(0.006) | 0.008***<br>(0.002) | 0.010*<br>(0.006) |
| WestF×Early | -0.052<br>(0.298) | 0.652**<br>(0.313) | 0.218***<br>(0.075) | 0.009<br>(0.006) | 0.006**<br>(0.003) | -0.013*<br>(0.007) | 0.010***<br>(0.002) | 0.005<br>(0.006) |
| F<sup>Early=Late</sup><br>p-value | 0.021<br>0.885 | 10.417<br>0.003 | 15.527<br>0.000 | 9.759<br>0.004 | 18.552<br>0.000 | 3.499<br>0.072 | 13.326<br>0.001 | 0.261<br>0.614 |
| F<sup>Early=Middle</sup><br>p-value | 1.331<br>0.258 | 7.733<br>0.010 | 7.049<br>0.013 | 6.428<br>0.017 | 1.878<br>0.181 | 2.222<br>0.147 | 2.638<br>0.116 | 1.324<br>0.260 |
| F<sup>Middle=Late</sup><br>p-value | 1.090<br>0.305 | 0.002<br>0.969 | 0.598<br>0.446 | 0.328<br>0.571 | 3.322<br>0.079 | 0.366<br>0.550 | 13.448<br>0.001 | 4.801<br>0.037 |
| Wild cluster bootstrap p-values: |  |  |  |  |  |  |  |  |
| WestF×Late | 0.530 | 0.102 | 0.140 | 0.078 | 0.062 | 0.824 | 0.729 | 0.742 |
| WestF×Middle | 0.303 | 0.586 | 0.914 | 0.410 | 0.837 | 0.687 | 0.001 | 0.094 |
| WestF×Early | 0.866 | 0.051 | 0.013 | 0.124 | 0.041 | 0.123 | 0.001 | 0.418 |
| Romano & Wolf p-values: |  |  |  |  |  |  |  |  |
| WestF×Late | 0.822 | 0.093 | 0.154 | 0.082 | 0.058 | 0.935 | 0.935 | 0.935 |
| WestF×Middle | 0.277 | 0.558 | 0.893 | 0.461 | 0.893 | 0.824 | 0.000 | 0.065 |
| WestF×Early | 0.949 | 0.034 | 0.006 | 0.071 | 0.034 | 0.048 | 0.000 | 0.325 |
| Observations | 42,826 | 42,826 | 42,826 | 42,826 | 42,826 | 42,826 | 42,826 | 42,826 |
| Controls and FE | ✓ | ✓ | ✓ | ✓ | ✓ | ✓ | ✓ | ✓ |
| Control Mean | 177.368 | 67.578 | 21.465 | 0.06 | 0.014 | 0.056 | 0.492 | 0.03 |

Note: Robust standard errors in parentheses clustered at the level of city. \*\*\* p<0.01, \*\* p<0.05, \* p<0.1. *Late* refers to births November 1944-January 1945, *Middle* to February 1945-April 1945, and *Early* to May 1945-July 1945. Controls include father's occupation, number of older brothers, birth order and religion. Fixed effects are included for city and for month of birth. Control Mean refers to the mean of the outcome for those born in the West Famine cities with postnatal exposure only (May-October 1944).

To examine whether any significant differences found in Table 3 might be driven by systematic differences between the specified groups other than by famine exposure, we then present in Table 4 the results from a placebo test, where we estimate the same equation as before but on the cohorts born two years later (i.e. between May 1946 and July 1947). As expected, we find no significant impacts in the placebo regressions for any of the outcomes. We then extend the placebo test to the sample used by Ravelli et al. (1976), which is twice as large, since it includes rural areas. The results are presented in the Appendix Table D2 and show that the placebo analysis fails: we find significant differences for weight, BMI, and underweight. This further confirms the importance of carefully selecting the control group, and casts doubts on the comparisons made in Ravelli et al. (1976).

**Table 4:**
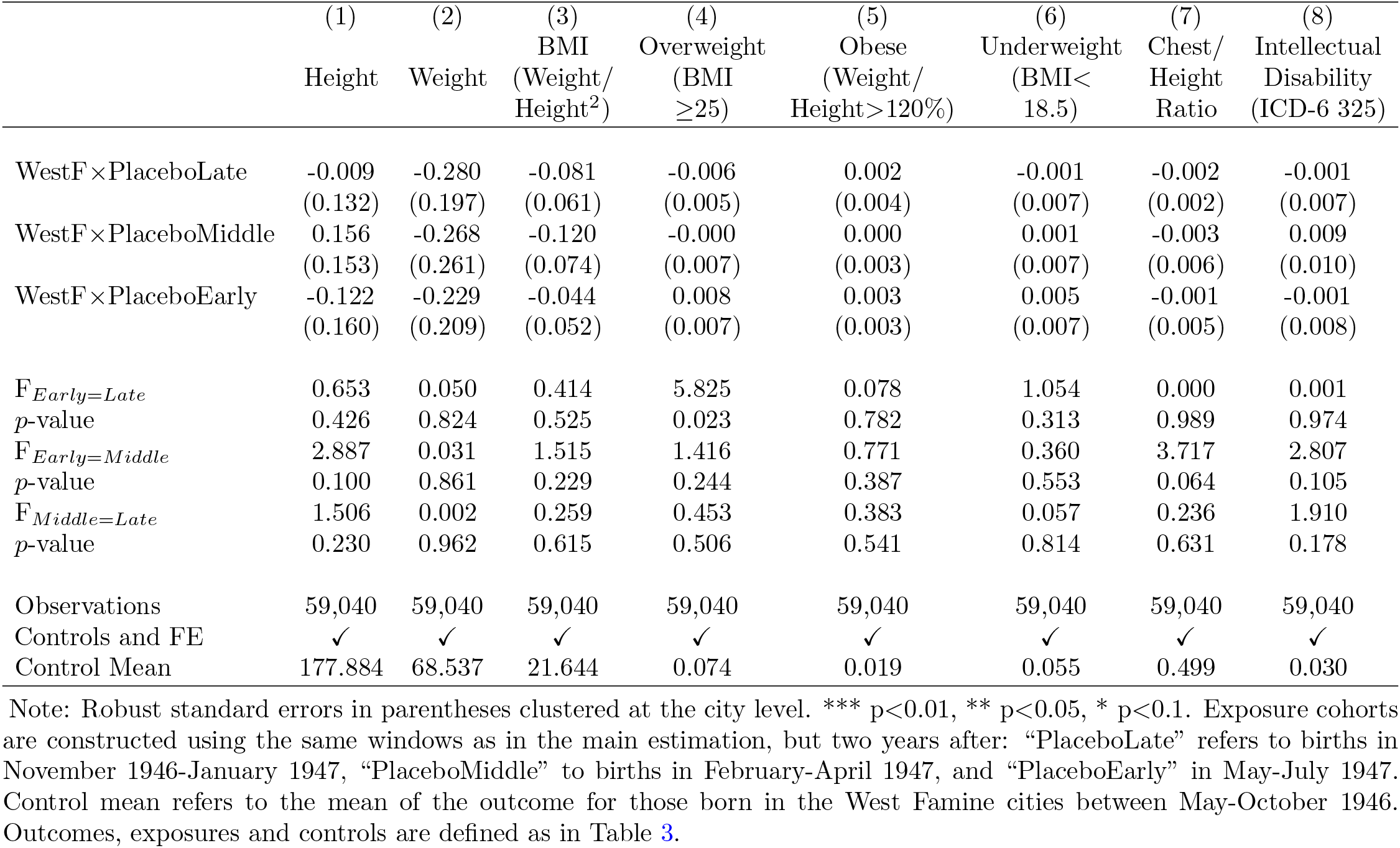
Placebo Difference-in-Differences Estimates of the Dutch Hunger Winter Effects on Age 18 Outcomes

Next, we combine our main wartime sample (i.e. births May 1944 - July 1945) and the placebo sample (births May 1946 - July 1947) in a triple difference specification. The results are reported in Table 5, where we display the triple interaction terms *WestF×Late×War*, *WestF×Middle×War*, and *WestF×Early×War* (from Equation 2). Most of our main results for exposure since the first trimester are confirmed, with the coefficients on the triple interaction terms increasing by almost 50% for weight, by 25% for BMI, and by 40% for being underweight; the coefficient on obesity, instead, is halved in size and no longer significant. Additionally, the other coefficients for exposure since middle and late gestation are no longer significant.^24^

**Table 5:**
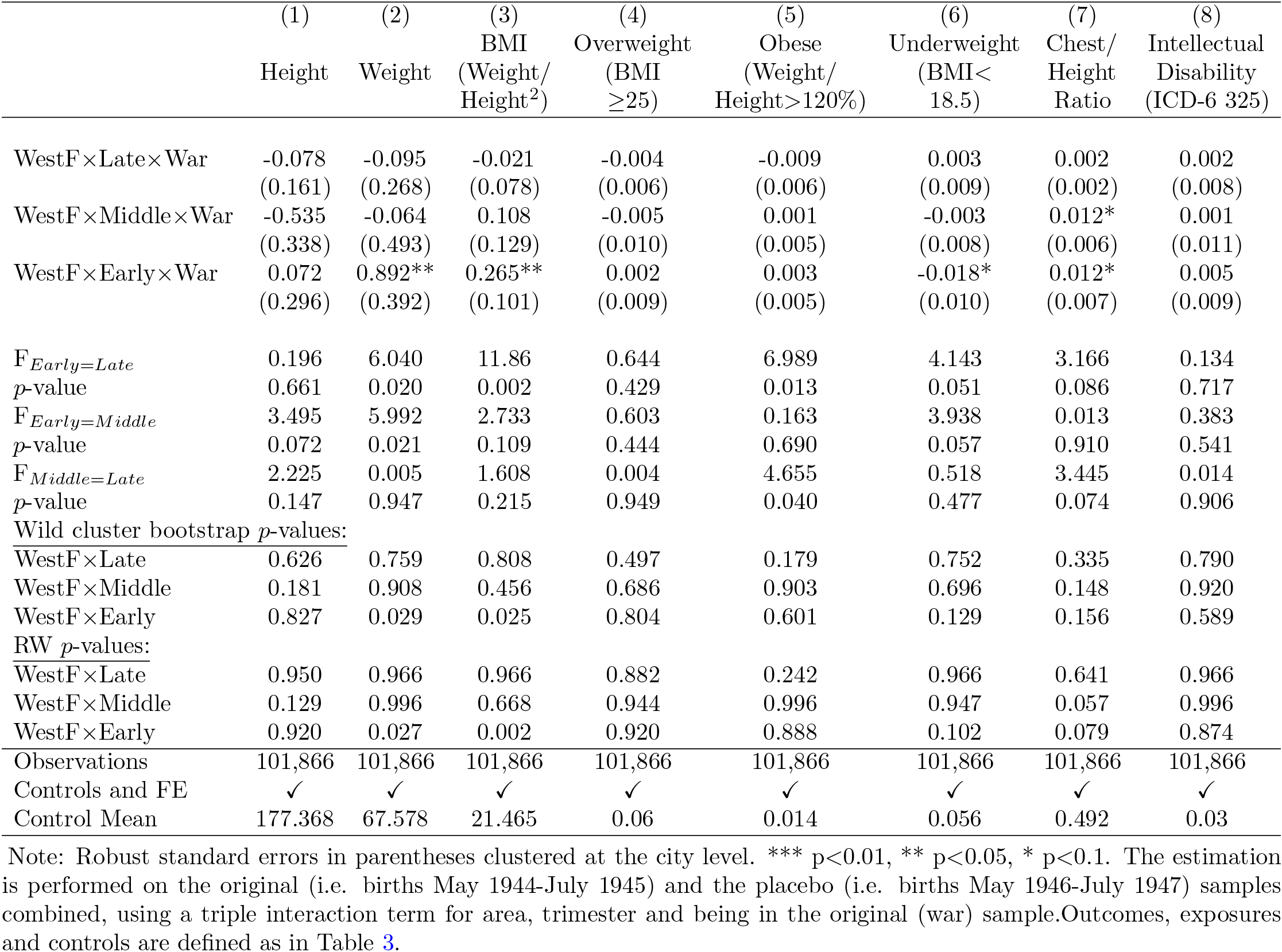
Triple Difference Estimates of the Dutch Hunger Winter Effects on Age 18 Outcomes

|  | (1) | (2) | (3) | (4) | (5) | (6) | (7) | (8) |
| --- | --- | --- | --- | --- | --- | --- | --- | --- |
|  | Height | Weight | BMI<br>(Weight/<br>Height<sup>2</sup>) | Overweight<br>(BMI<br>≥25) | Obese<br>(Weight/<br>Height>120%) | Underweight<br>(BMI<<br>18.5) | Chest/<br>Height<br>Ratio | Intellectual<br>Disability<br>(ICD-6 325) |
| WestF×Late×War | -0.078<br>(0.161) | -0.095<br>(0.268) | -0.021<br>(0.078) | -0.004<br>(0.006) | -0.009<br>(0.006) | 0.003<br>(0.009) | 0.002<br>(0.002) | 0.002<br>(0.008) |
| WestF×Middle×War | -0.535<br>(0.338) | -0.064<br>(0.493) | 0.108<br>(0.129) | -0.005<br>(0.010) | 0.001<br>(0.005) | -0.003<br>(0.008) | 0.012*<br>(0.006) | 0.001<br>(0.011) |
| WestF×Early×War | 0.072<br>(0.296) | 0.892**<br>(0.392) | 0.265**<br>(0.101) | 0.002<br>(0.009) | 0.003<br>(0.005) | -0.018*<br>(0.010) | 0.012*<br>(0.007) | 0.005<br>(0.009) |
| $F^{Early=Late}$<br>$p$-value | 0.196<br>0.661 | 6.040<br>0.020 | 11.86<br>0.002 | 0.644<br>0.429 | 6.989<br>0.013 | 4.143<br>0.051 | 3.166<br>0.086 | 0.134<br>0.717 |
| $F^{Early=Middle}$<br>$p$-value | 3.495<br>0.072 | 5.992<br>0.021 | 2.733<br>0.109 | 0.603<br>0.444 | 0.163<br>0.690 | 3.938<br>0.057 | 0.013<br>0.910 | 0.383<br>0.541 |
| $F^{Middle=Late}$<br>$p$-value | 2.225<br>0.147 | 0.005<br>0.947 | 1.608<br>0.215 | 0.004<br>0.949 | 4.655<br>0.040 | 0.518<br>0.477 | 3.445<br>0.074 | 0.014<br>0.906 |
| Wild cluster bootstrap $p$-values: | | | | | | | | |
| WestF×Late | 0.626 | 0.759 | 0.808 | 0.497 | 0.179 | 0.752 | 0.335 | 0.790 |
| WestF×Middle | 0.181 | 0.908 | 0.456 | 0.686 | 0.903 | 0.696 | 0.148 | 0.920 |
| WestF×Early | 0.827 | 0.029 | 0.025 | 0.804 | 0.601 | 0.129 | 0.156 | 0.589 |
| RW $p$-values: | | | | | | | | |
| WestF×Late | 0.950 | 0.966 | 0.966 | 0.882 | 0.242 | 0.966 | 0.641 | 0.966 |
| WestF×Middle | 0.129 | 0.996 | 0.668 | 0.944 | 0.996 | 0.947 | 0.057 | 0.996 |
| WestF×Early | 0.920 | 0.027 | 0.002 | 0.920 | 0.888 | 0.102 | 0.079 | 0.874 |
| Observations | 101,866 | 101,866 | 101,866 | 101,866 | 101,866 | 101,866 | 101,866 | 101,866 |
| Controls and FE | ✓ | ✓ | ✓ | ✓ | ✓ | ✓ | ✓ | ✓ |
| Control Mean | 177.368 | 67.578 | 21.465 | 0.06 | 0.014 | 0.056 | 0.492 | 0.03 |

Note: Robust standard errors in parentheses clustered at the city level. \*\*\* $p < 0.01$ , \*\* $p < 0.05$ , \* $p < 0.1$ . The estimation is performed on the original (i.e. births May 1944-July 1945) and the placebo (i.e. births May 1946-July 1947) samples combined, using a triple interaction term for area, trimester and being in the original (war) sample. Outcomes, exposures and controls are defined as in Table 3.

Lastly, we repeated the estimations in Tables 3 and 5 by excluding one control region at a time to test whether the results are sensitive to the choice of the comparison group. Results are presented in Figures C1 and C2 in the Appendix, where the plots show the interaction term estimates as in the Tables 3 and 5, along with their 90% confidence intervals. Across both models, the BMI results are robust to the choice of the region used as comparison group.

### 5.2 Mechanisms

We now exploit our newly digitised data to investigate the mechanisms through which being in utero during the Dutch Hunger Winter led to adverse outcomes at 18; we focus on the outcomes for which we have found a significant effect in the main specification (equation 1). Table 6 shows the results from our basic difference-in-differences specification, with the addition of variables on calories, protein share, warfare deaths, temperature and weeks since liberation, separately by trimester.^25^

**Table 6:** Difference-in-Differences Estimates and Mechanisms of the Dutch Hunger Winter Effects on Age 18 Outcomes

|  | (1)<br>Weight | (2)<br>BMI<br>(Weight/<br>Height<sup>2</sup>) | (3)<br>Overweight<br>(BMI<br>≥25) | (4)<br>Obese<br>(Weight/<br>Height>120%) | (5)<br>Underweight<br>(BMI<<br>18.5) | (6)<br>Chest/<br>Height<br>Ratio |
| --- | --- | --- | --- | --- | --- | --- |
| WestF×Late | -0.382<br>(0.247) | -0.037<br>(0.071) | -0.005<br>(0.006) | -0.004<br>(0.003) | 0.001<br>(0.008) | -0.002<br>(0.002) |
| WestF×Middle | -0.426<br>(0.362) | 0.009<br>(0.101) | 0.000<br>(0.009) | 0.007*<br>(0.004) | 0.001<br>(0.010) | 0.003<br>(0.004) |
| WestF×Early | 0.482<br>(0.319) | 0.141<br>(0.107) | 0.010<br>(0.009) | 0.007<br>(0.005) | -0.002<br>(0.012) | 0.004<br>(0.004) |
| log(Warfare1stTR+1) | 0.040<br>(0.056) | -0.010<br>(0.011) | 0.001<br>(0.001) | 0.001<br>(0.001) | 0.002<br>(0.001) | 0.000<br>(0.000) |
| log(Warfare2ndTR+1) | 0.031<br>(0.054) | 0.013<br>(0.014) | 0.003**<br>(0.001) | -0.000<br>(0.001) | -0.001<br>(0.001) | -0.001<br>(0.000) |
| log(Warfare3rdTR+1) | 0.100*<br>(0.053) | 0.030**<br>(0.013) | 0.001<br>(0.001) | 0.001<br>(0.001) | -0.001<br>(0.001) | 0.000<br>(0.000) |
| -Calories1stTR | 0.575<br>(0.972) | -0.059<br>(0.245) | 0.034<br>(0.022) | 0.013<br>(0.015) | 0.039<br>(0.029) | 0.006<br>(0.005) |
| -ProteinShare1stTR | -0.061<br>(0.352) | 0.011<br>(0.079) | -0.001<br>(0.008) | 0.006<br>(0.005) | 0.008<br>(0.008) | 0.001<br>(0.001) |
| -Calories2ndTR | -0.773<br>(0.898) | -0.086<br>(0.208) | -0.031*<br>(0.016) | -0.021<br>(0.014) | -0.026<br>(0.016) | -0.003<br>(0.003) |
| -ProteinShare2ndTR | 0.171<br>(0.145) | 0.041<br>(0.040) | -0.001<br>(0.004) | 0.002<br>(0.003) | -0.006<br>(0.005) | 0.001<br>(0.001) |
| -Calories3rdTR | -0.839<br>(0.602) | -0.298<br>(0.201) | -0.014<br>(0.015) | -0.003<br>(0.006) | 0.029**<br>(0.011) | 0.005<br>(0.004) |
| -ProteinShare3rdTR | 0.428***<br>(0.115) | 0.105***<br>(0.023) | 0.008**<br>(0.003) | 0.003*<br>(0.002) | -0.003<br>(0.003) | 0.001*<br>(0.000) |
| Wild cluster bootstrap *p*-values: |  |  |  |  |  |  |
| WestF×Late | 0.144 | 0.614 | 0.441 | 0.208 | 0.936 | 0.352 |
| WestF×Middle | 0.282 | 0.937 | 0.969 | 0.127 | 0.916 | 0.592 |
| WestF×Early | 0.165 | 0.266 | 0.302 | 0.243 | 0.906 | 0.395 |
| log(Warfare1stTR+1) | 0.530 | 0.340 | 0.565 | 0.463 | 0.215 | 0.814 |
| log(Warfare2ndTR+1) | 0.584 | 0.335 | 0.080 | 0.479 | 0.398 | 0.225 |
| log(Warfare3rdTR+1) | 0.170 | 0.111 | 0.713 | 0.408 | 0.320 | 0.812 |
| -Calories1stTR | 0.577 | 0.830 | 0.135 | 0.401 | 0.200 | 0.331 |
| -ProteinShare1stTR | 0.879 | 0.902 | 0.922 | 0.351 | 0.356 | 0.624 |
| -Calories2ndTR | 0.427 | 0.690 | 0.109 | 0.180 | 0.100 | 0.441 |
| -ProteinShare2ndTR | 0.260 | 0.341 | 0.838 | 0.482 | 0.274 | 0.330 |
| -Calories3rdTR | 0.214 | 0.206 | 0.393 | 0.667 | 0.017 | 0.225 |
| -ProteinShare3rdTR | 0.003 | 0.001 | 0.046 | 0.087 | 0.287 | 0.067 |
| RW *p*-values: |  |  |  |  |  |  |
| WestF×Late | 0.203 | 0.702 | 0.551 | 0.294 | 0.915 | 0.410 |
| WestF×Middle | 0.339 | 0.997 | 0.997 | 0.109 | 0.997 | 0.719 |
| WestF×Early | 0.218 | 0.295 | 0.295 | 0.295 | 0.848 | 0.295 |
| log(Warfare1stTR+1) | 0.636 | 0.601 | 0.636 | 0.636 | 0.360 | 0.653 |
| log(Warfare2ndTR+1) | 0.633 | 0.585 | 0.065 | 0.633 | 0.623 | 0.291 |
| log(Warfare3rdTR+1) | 0.078 | 0.030 | 0.774 | 0.489 | 0.489 | 0.774 |
| -Calories1stTR | 0.561 | 0.721 | 0.170 | 0.487 | 0.230 | 0.359 |
| -ProteinShare1stTR | 0.984 | 0.984 | 0.984 | 0.421 | 0.532 | 0.822 |
| -Calories2ndTR | 0.373 | 0.573 | 0.082 | 0.098 | 0.098 | 0.373 |
| -ProteinShare2ndTR | 0.387 | 0.387 | 0.796 | 0.461 | 0.387 | 0.387 |
| -Calories3rdTR | 0.154 | 0.140 | 0.372 | 0.581 | 0.010 | 0.154 |
| -ProteinShare3rdTR | 0.000 | 0.000 | 0.012 | 0.052 | 0.174 | 0.052 |
| Observations | 40,950 | 40,950 | 40,950 | 40,950 | 40,950 | 40,950 |
| Controls and FE | ✓ | ✓ | ✓ | ✓ | ✓ | ✓ |

Note: Robust standard errors in parentheses clustered at the level of city. \*\*\* $p < 0.01$ , \*\* $p < 0.05$ , \* $p < 0.1$ . Controls include father's occupation, number of older brothers, birth order, religion, temperature by trimester and weeks since liberation for the South. FE are included for city and for month of birth. Outcome and *Early*, *Middle*, *Late* definitions as in Table 3. Calories are in thousands, see Table 2 for summary statistics.

Our results show that the stress and nutritional channels play a significant role in explaining the effects of being born during the famine on health at age 18. In particular, the impacts on weight and BMI (whose coefficients are reduced by 26% and 35%, respectively) for those exposed since early gestation appear to be driven by a combination of increased exposure to warfare and reduction of proteins in the rations (irrespective of the caloric content), which became particularly severe at the end of the famine, when those with early exposure were in the third trimester. The impact on chest-height ratio (whose coefficient is reduced by 60%) appears entirely driven by a reduction in protein share at the end of gestation, which also has a significant effect on the prevalence of overweight and obesity at 18. Note the coefficients on overweight and obesity are unchanged in size as compared to Table 3, but they are less precisely estimated.^26^ These results are robust to the choice of cities included in our sample (see Figure C3 in the Appendix).

These findings are consistent with an established nutrition literature which has linked, in animal models, a protein-restricted diet in mothers with impaired metabolic function in the offspring (Stocker et al., 2005); the effects of unbalanced protein intake in pregnancy have also been found in humans (see e.g. Imdad and Bhutta (2011) for a review of studies on birth outcomes. Additionally, the cohorts exposed since earlier in gestation have also experienced a ‘mismatch’ between a harsher prenatal famine environment and a more prosperous post-liberation environment, which could have exacerbated the effects of in utero shocks (Gluckman and Hanson, 2004). However, while we are able to disentangle key factors correlated with early in utero exposure to the Dutch Hunger Winter, we are unable to account for other possible channels, such as negative income shocks, possible exposure to toxins, and behavioural responses:^27^ hence, our reduced-form coefficients might be bigger or smaller than a ‘purely biological effect’ (Yi et al., 2015).

### 5.3 Birth Outcomes

As described in Section 3.3, to explore impacts of famine exposure on birth outcomes, we then use the additional dataset with information collected in selected birth hospital clinics in three treated and two control cities. For consistency with the recruits data, we focus on the sample of male newborns. We first show in Table D4 that we are able to replicate the main results in the subsample of cities for which we have the hospital birth data. We then present the birth results in Table 7. They show a reduction in birth weight and placenta weight for those exposed since middle and early gestation, respectively; however, none of the effects are robust to controlling for multiple hypothesis testing. In addition, we find no effect on gestational age (column 7) and on sex ratio (column 8, where we use all births, i.e. males and females) for any of the exposure groups.^28^ The fact that there are no effects on sex ratio and gestational age further validates our strategy of using the date of birth to identify the date of conception (and so the exposure by trimester), and rules out any concerns related to the fact that we don’t observe miscarriages in our mortality data.

**Table 7:** Difference-in-Differences Results for the Birth Outcomes

|  | (1)<br>Birth<br>Weight | (2)<br>Low Birth<br>Weight | (3)<br>Birth<br>Length | (4)<br>Head<br>Circumference | (5)<br>Placenta<br>Weight | (6)<br>Gestational<br>Age | (7)<br>Male |
| --- | --- | --- | --- | --- | --- | --- | --- |
| West × Late | -76.681<br>(87.265) | -0.001<br>(0.034) | 0.010<br>(0.372) | 0.887*<br>(0.508) | -38.839<br>(26.568) | -0.079<br>(0.286) | 0.030<br>(0.057) |
| West × Middle | -168.993**<br>(82.816) | -0.007<br>(0.036) | -0.522<br>(0.381) | 0.250<br>(0.411) | -39.760*<br>(22.748) | 0.050<br>(0.299) | 0.052<br>(0.052) |
| West × Early | -76.137<br>(78.591) | -0.008<br>(0.033) | -0.320<br>(0.379) | 0.181<br>(0.409) | -43.531*<br>(23.034) | -0.023<br>(0.298) | 0.004<br>(0.053) |
| $F_{Early=Late}$ | 0.001 | 0.058 | 1.137 | 2.507 | 0.043 | 0.036 | 0.243 |
| $p$-value | 0.994 | 0.809 | 0.286 | 0.114 | 0.836 | 0.849 | 0.622 |
| $F_{Early=Middle}$ | 1.981 | 0.002 | 0.470 | 0.055 | 0.048 | 0.058 | 1.117 |
| $p$-value | 0.159 | 0.962 | 0.493 | 0.815 | 0.826 | 0.809 | 0.291 |
| $F_{Middle=Late}$ | 1.377 | 0.030 | 2.983 | 2.025 | 0.002 | 0.195 | 0.184 |
| $p$-value | 0.241 | 0.862 | 0.084 | 0.155 | 0.967 | 0.658 | 0.668 |
| RW $p$-values: | | | | | | | |
| West × Late | 0.785 | 0.988 | 0.988 | 0.376 | 0.482 | 0.988 | |
| West × Middle | 0.183 | 0.964 | 0.514 | 0.896 | 0.324 | 0.964 | |
| West × Early | 0.809 | 0.957 | 0.865 | 0.957 | 0.248 | 0.957 | |
| Observations | 1,931 | 1,931 | 1,869 | 1,275 | 1,295 | 1,259 | 3,697 |
| Control and FE | ✓ | ✓ | ✓ | ✓ | ✓ | ✓ | ✓ |
| Control Mean | 3,414.378 | 0.072 | 50.584 | 38.857 | 633.051 | 39.518 | 0.531 |

Note: Columns (1)-(6) are based on males only sample. Column (7) sample includes all births (males and females). Robust standard errors in parentheses. \*\*\* $p < 0.01$ , \*\* $p < 0.05$ , \* $p < 0.1$ . All models include mother's age and FE for city and for month of birth. Control Mean refers to the mean of the outcome for those born in the West Famine area with postnatal exposure only. Results from hospital records in the birth data. *Early*, *Middle*, *Late* definitions as in Table 3.

### 5.4 Accounting for Selection

In this last section we account for selective mortality using a copula-based approach. Recall that our main analysis is based on military recruits data, which was collected when the respondents were 18 years old, conditional on being alive and resident in the Netherlands; additionally, since the cohorts born in the Western cities faced much worse conditions than those in the rest of the country, we expect them to be less likely to survive. While the vast majority of the literature on the developmental origins of health does not account for survivorship bias, we are able to overcome this limitation, using the additional data assembled as explained in section 3.2. These data are displayed in Figure D2, which shows that there is substantial variation in survival rates (proportion of those born in the period May 1944-July 1945 in one of the selected cities who are alive at age 18) across cohorts and cities. Table D5 shows the results for the selection equation: the individuals exposed during gestation have indeed a lower probability of surviving until age 18 as compared to those with only postnatal exposure, with the greater reduction in survival experienced by those exposed since mid-gestation. Reassuringly, the excluded variables have the expected sign: a greater availability of medical staff in the city and a larger number of religious groups is associated with greater survival, while living in a larger municipality is associated with lower survival. Consistently with our Monte Carlo exercise, the results are robust to using different subsets of the exclusion restrictions.

Additionally, we experiment with a series of Monte Carlo simulations to compare alternative estimation methods to account for the problem of selective survival (see Appendix E), and understand their relative performance in a setup similar to ours. In doing so, we extend previous research on selection bias in similar setups which has examined selection models with normal error structures (Grilli and Rampichini, 2010), by relaxing the assumption of bivariate normality with the use of copulas, that preserve the dependence structure and allow various forms of excess joint asymmetry, skewness or kurtosis.^29^ The setup of the simulation design includes a cluster-level selection equation and an individual-level outcome equation, both with a cluster-level variable (*f_j_*, i.e. famine incidence), individual-level variables (*x_ij_*), a cluster-level error (*u_j_*) and an individual-level error (*ε_ij_*); additionally, the selection equation includes a regional-level variable (*z_j_*) as exclusion restriction to facilitate identification.

Our simulation study shows that the best performing estimation technique depends on the crucial assumption on the correlation of the error terms among the outcome (indexed with 1) and selection (indexed with 2) equations at each level (i.e. between *u*_1_*_j_* and *u*_2_*_j_*, and *ε*_1_*_ij_* and *ε*_2_*_ij_*). If a correlation exists only at regional level (thus *Cov*(*ε*_1_*_ij_, ε*_2_*_ij_*) = 0), one can use Inverse Probability Weighting, or a GLS random-effects model to successfully account for the selective survival (Table E1). On the other hand, if a correlation also exists at the individual level (thus *Cov*(*u*_1_*_j_, u*_2_*_j_*) = 0), both approaches will yield inconsistent estimates; however, in this case a selection model will produce consistent estimates. Table E1 further shows that a Heckman-type selection model performs marginally better than a copula-based one in the presence of a “strong” exclusion restriction, and definitely worse in case of a “weak” exclusion and error correlation at both cluster level and individual level; in general, however, the copula-based selection model is always the most efficient. This reassures us on the use of a copula-based approach to account for selective survival in estimating the impact of the famine on adolescent health.

We then use the new data on the survival probability to estimate models which account for sample selection departing from the standard assumption of bivariate normality of the errors by using more flexible copulas functions. The results are presented in Table 8, and show evidence of *both* selection *and* scarring effects.^30^ First of all, as expected, the survivors appear to be positively selected: the sign and magnitude of Kendal’s *τ* ^31^ show that the survivors are less likely to be either underweight or overweight, and have on average a taller height and larger weight, and higher BMI; in other words, those taller and more robust (without being overweight) appear to have been more likely to survive, and so the BMI distribution of the survivors appears truncated both on the left and the right.^32^ Second, once we account for survival, we still detect a significant scarring effect, with those exposed since early gestation having a higher BMI and CHR (by 0.188 and 0.011) and being 0.7 p.p. and 0.5 p.p. more likely to be overweight and obese and 0.8 p.p. less likely to be underweight than the controls, respectively – all impacts in line with the baseline specification (Table 3), as expected given small selection effects among those with early exposure (Table D5). Third, accounting for survival seems particularly important to understand the famine impacts on intellectual disability (column 8): we detect not only a negative selection effect, but also a significant scarring effect of famine exposure in middle and late gestation on intellectual disability, by 1.4 p.p. and 0.7 p.p., respectively (w.r.t. a control mean of 3%).

**Table 8:**
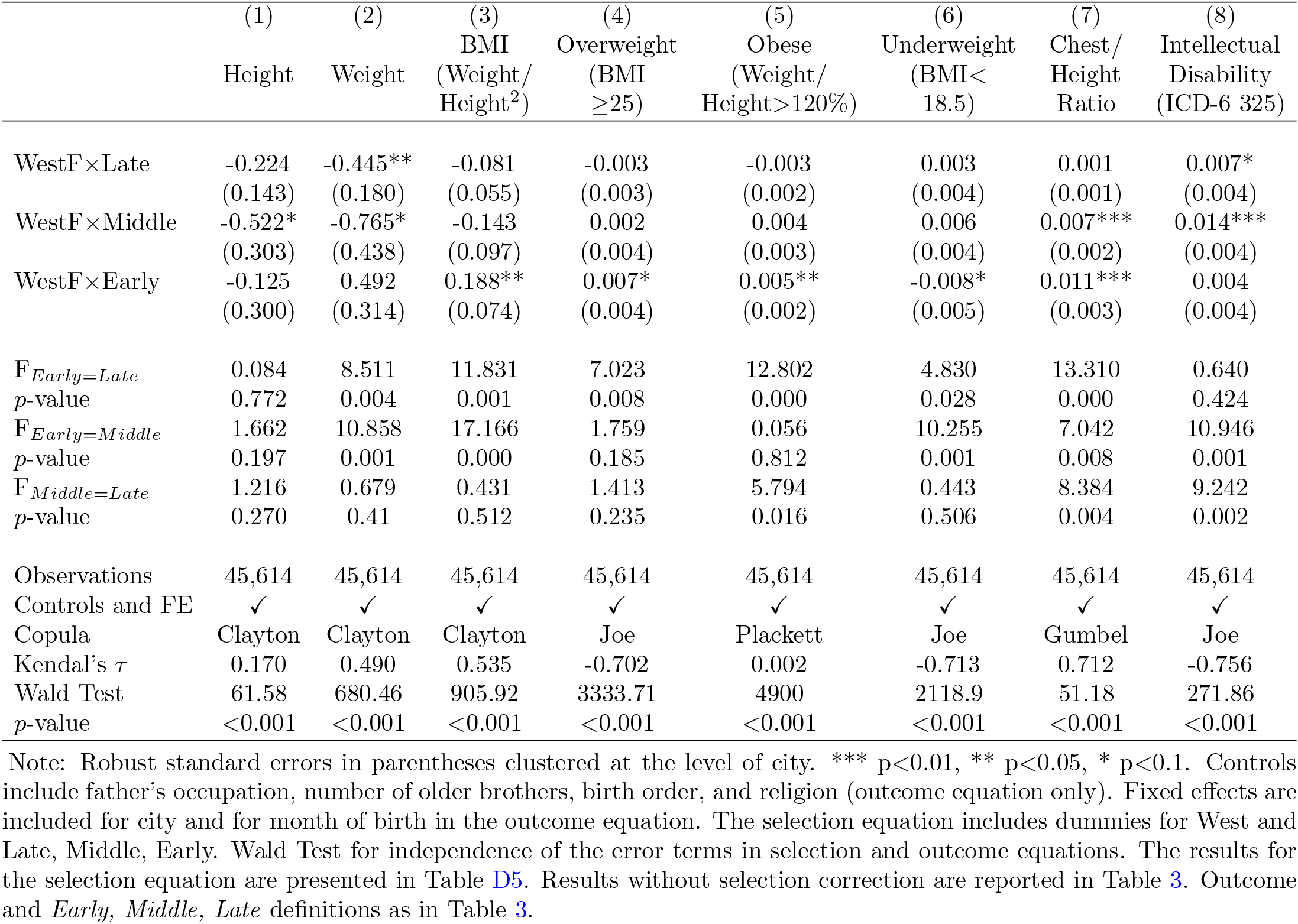
Difference-in-Differences Estimates of the Dutch Hunger Winter Effects on Age 18 Outcomes with Sample Selection Correction using Copulas

In sum, also after accounting for survivor bias, we find that prenatal exposure to the famine caused increases in both physical health conditions (including both overweight and obesity) and intellectual disabilities.

## 6 Conclusion

In this paper, we have investigated impacts, mechanisms and selection effects of prenatal exposure to multiple shocks, by exploiting the unique natural experiment of the Dutch Hunger Winter, which hit abruptly the Western Netherlands at the end of World War II. We have linked military recruits data on the cohorts born in the years surrounding World War II, used in landmark epidemiological papers, with newly digitised historical records on calories and nutrient composition of the war rations, daily temperature, and warfare deaths. We have also collected a variety of demographic and health indicators for all the cities in the Netherlands since the mid 1930s, to carefully select the cities in the control group, on the basis of common trends. Armed with this new rich resource, we have provided several contributions to the interdisciplinary literature on the developmental origins of health.

First, we have confirmed previous epidemiological findings that the cohorts exposed to the Dutch Hunger Winter since early gestation have an increased probability of being overweight at age 18, and we have provided new parallel findings on chest/height ratio. Second, by providing different placebo and robustness tests, we have shown that the careful selection of cities for the control group matters. Third, we have found that the adverse health effects of early in utero exposure to the Dutch Hunger Winter are in part explained by a combination of warfare exposure and protein reduction in the rations at the end of gestation, while the cold winter temperature does not seem to play a role. Fourth, we have carried out a novel selection correction by means of copula models (supported by a Monte Carlo investigation), and we have shown evidence of both selection (‘survival of the fittest’) and scarring effects; crucially, even after accounting for selective survival, early in utero exposure to the Dutch Hunger Winter leads to long-term scarring in physical health and mental capability.

Our work has advanced our understanding of the impacts of one of the best-studied early life shocks, and employed an empirical approach which can be used to account for selection in studies with a similar setup. Although our study is based on a historical context, wars and famines are still a reality in the modern world, so that lessons learnt from the Dutch Hunger Winter can still be applied today.

## Data Availability

Please contact the authors for data availability.

## Online Appendix

### Appendix A Historical documents

**Figure A1:**
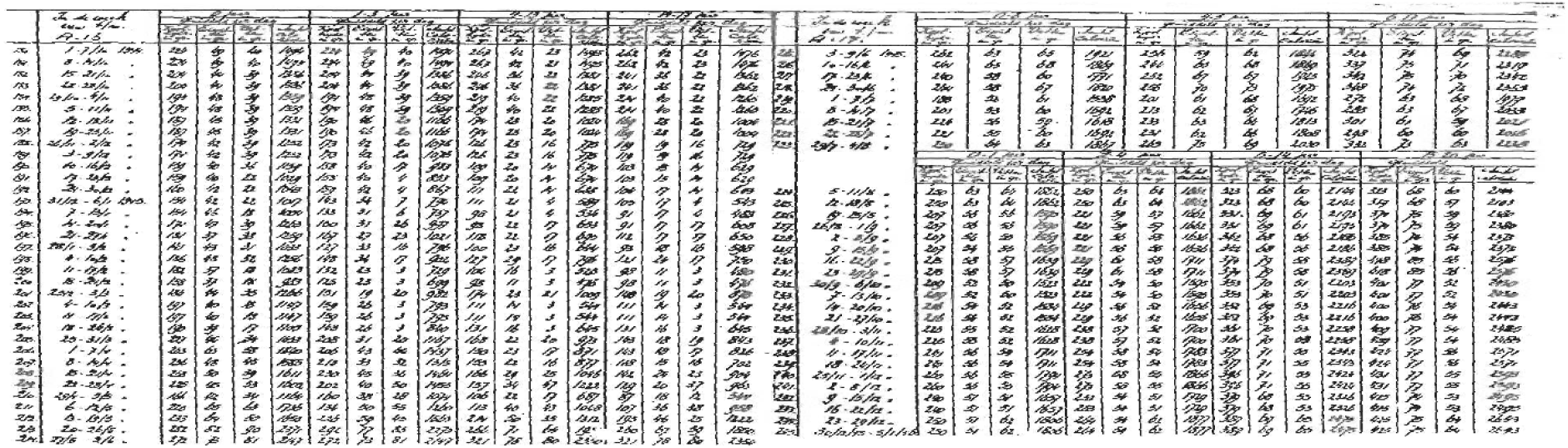
Rations on calories and nutrients composition Note: Calories and nutrients composition of the rations from the official war information on the rations (Departement van Landbouw en Visserij, 1946).

### Appendix B Pretrends for selection of cities

**Figure B1:**
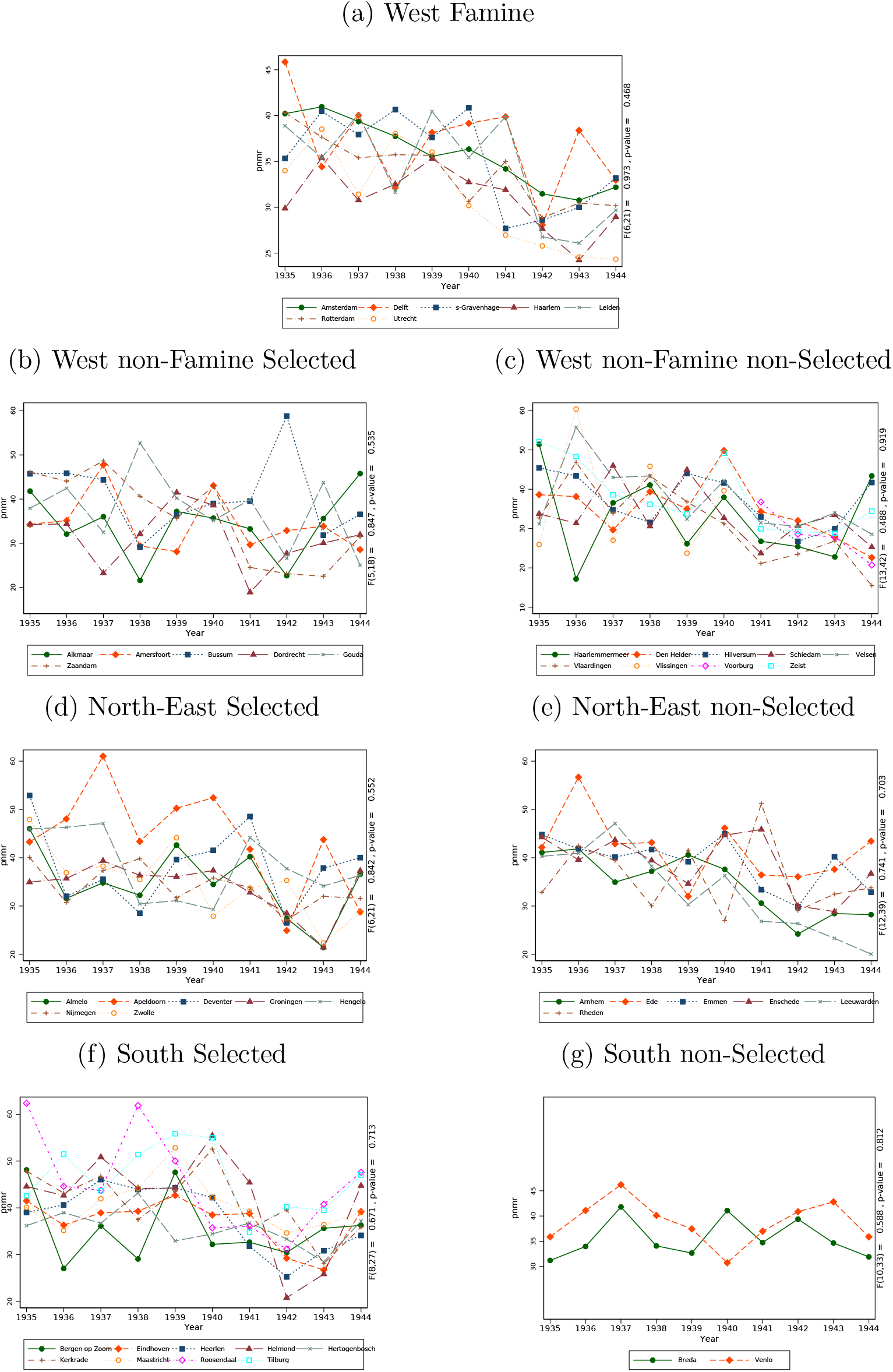
Pretrends for Postnatal Mortality Rate Note: Pretrends shown for groups of selected, and non-selected, in the analysis cities. A Wald test is performed in each group, by fitting a linear regression with city-specific slopes and testing jointly whether the slopes are equal.

**Figure B2:**
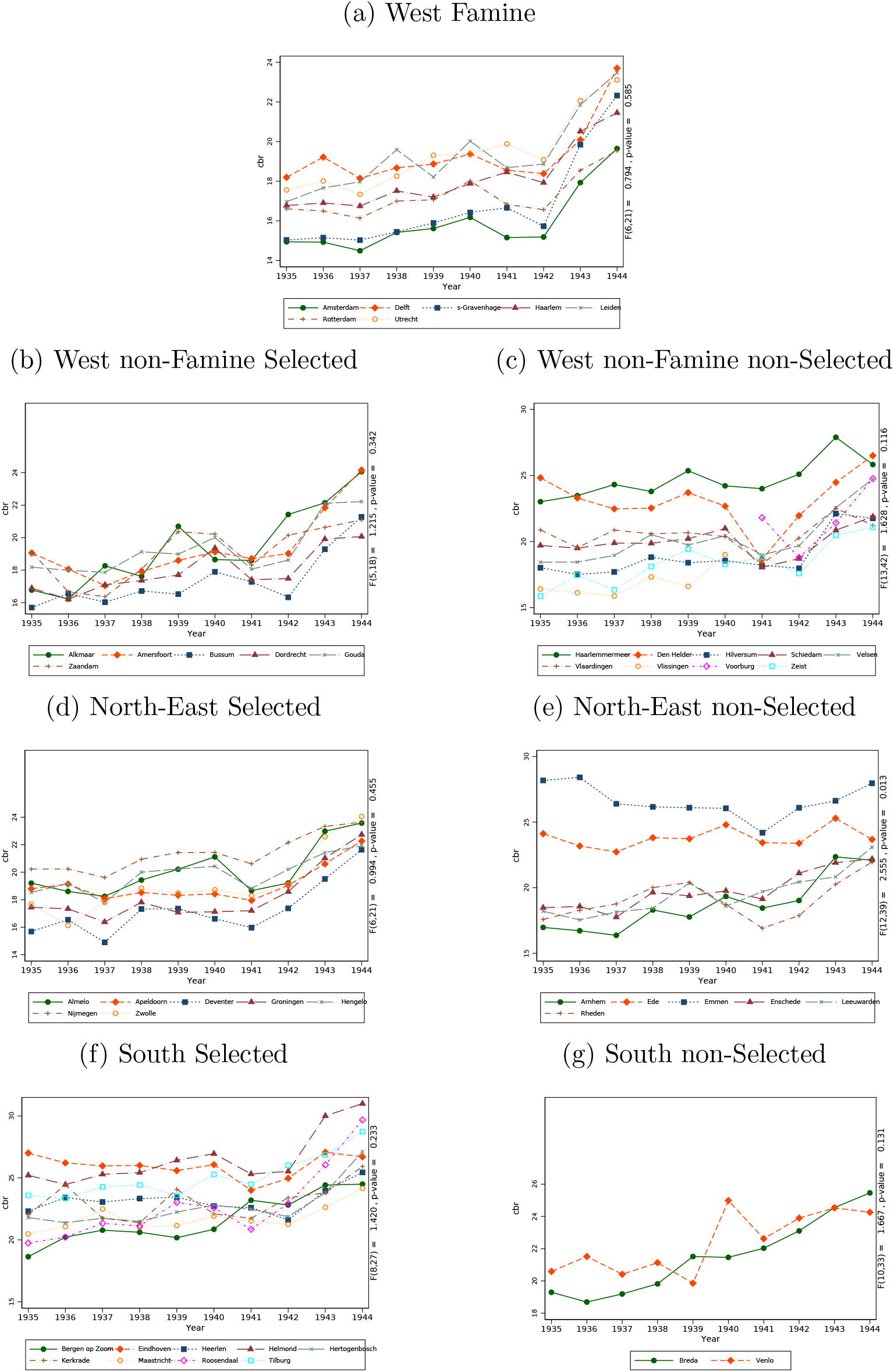
Pretrends for Crude Birth Rate Note: Pretrends shown for groups of selected, and non-selected, in the analysis cities. A Wald test is performed in each group, by fitting a linear regression with city-specific slopes and testing jointly whether the slopes are equal.

**Figure B3:**
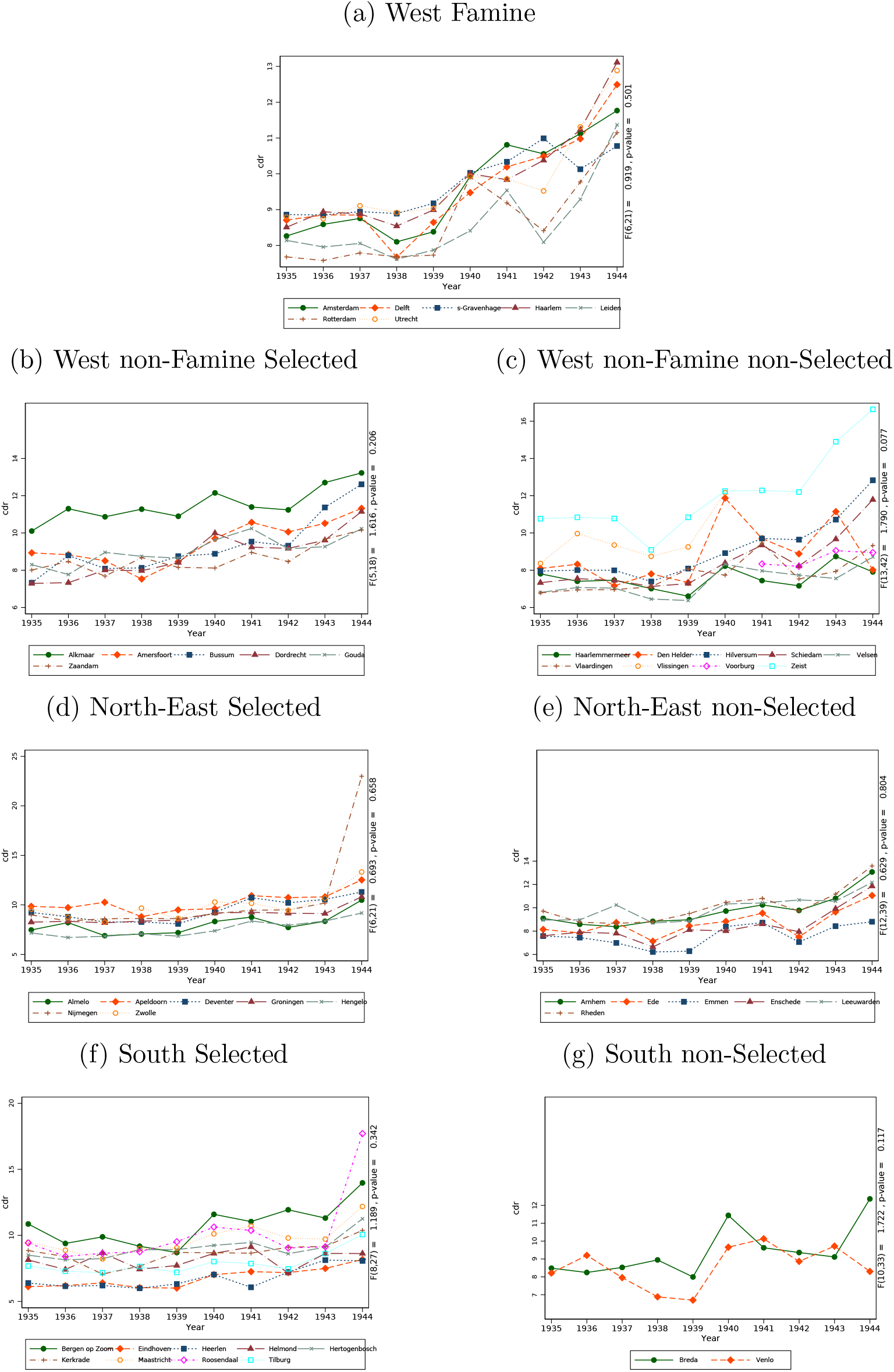
Pretrends for Crude Death Rate Note: Pretrends shown for groups of selected, and non-selected, in the analysis cities. A Wald test is performed in each group, by fitting a linear regression with city-specific slopes and testing jointly whether the slopes are equal.

**Figure B4:**
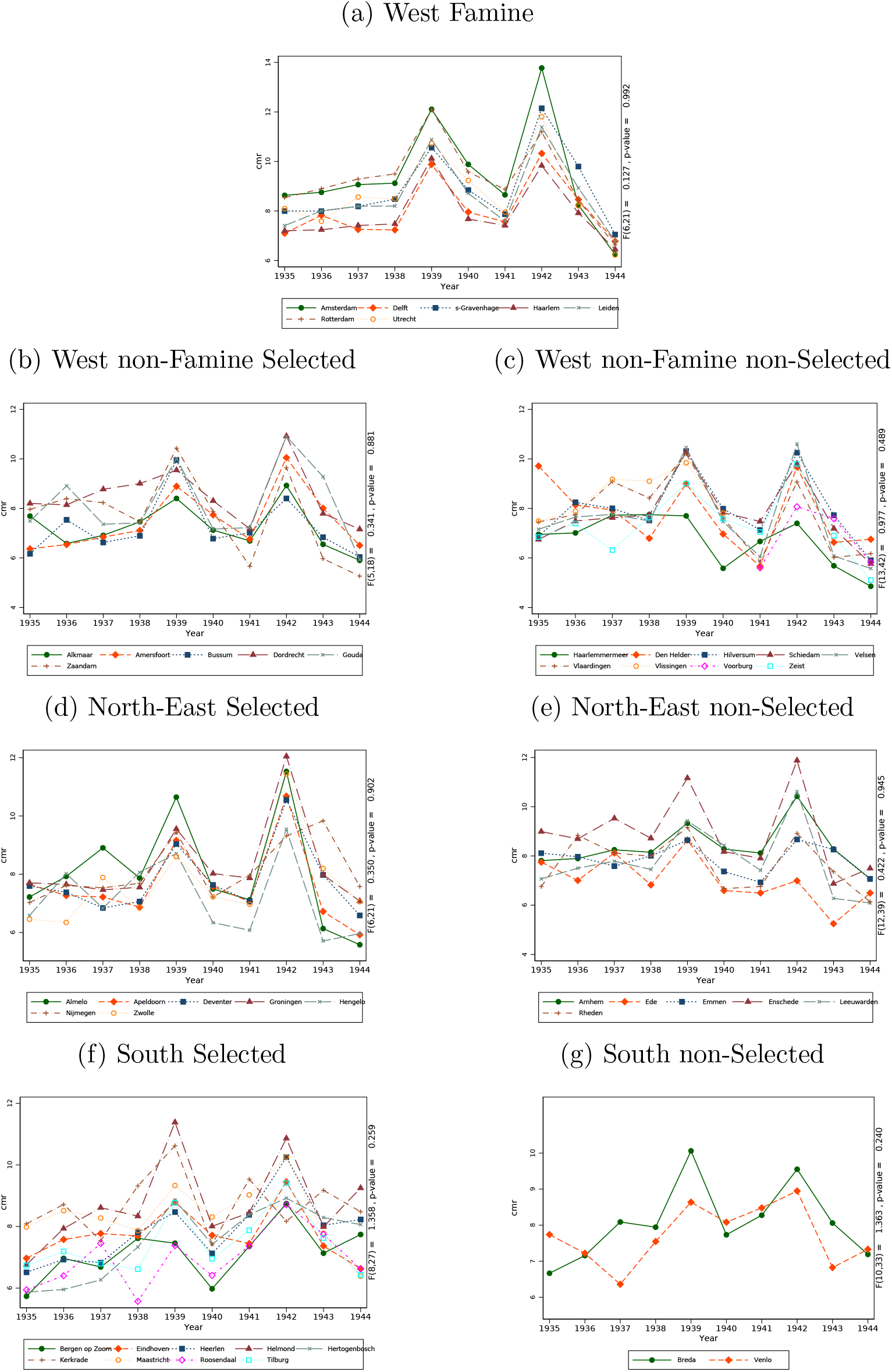
Pretrends for Crude Marriage Rate Note: Pretrends shown for groups of selected, and non-selected, in the analysis cities. A Wald test is performed in each group, by fitting a linear regression with city-specific slopes and testing jointly whether the slopes are equal.

**Figure B5:**
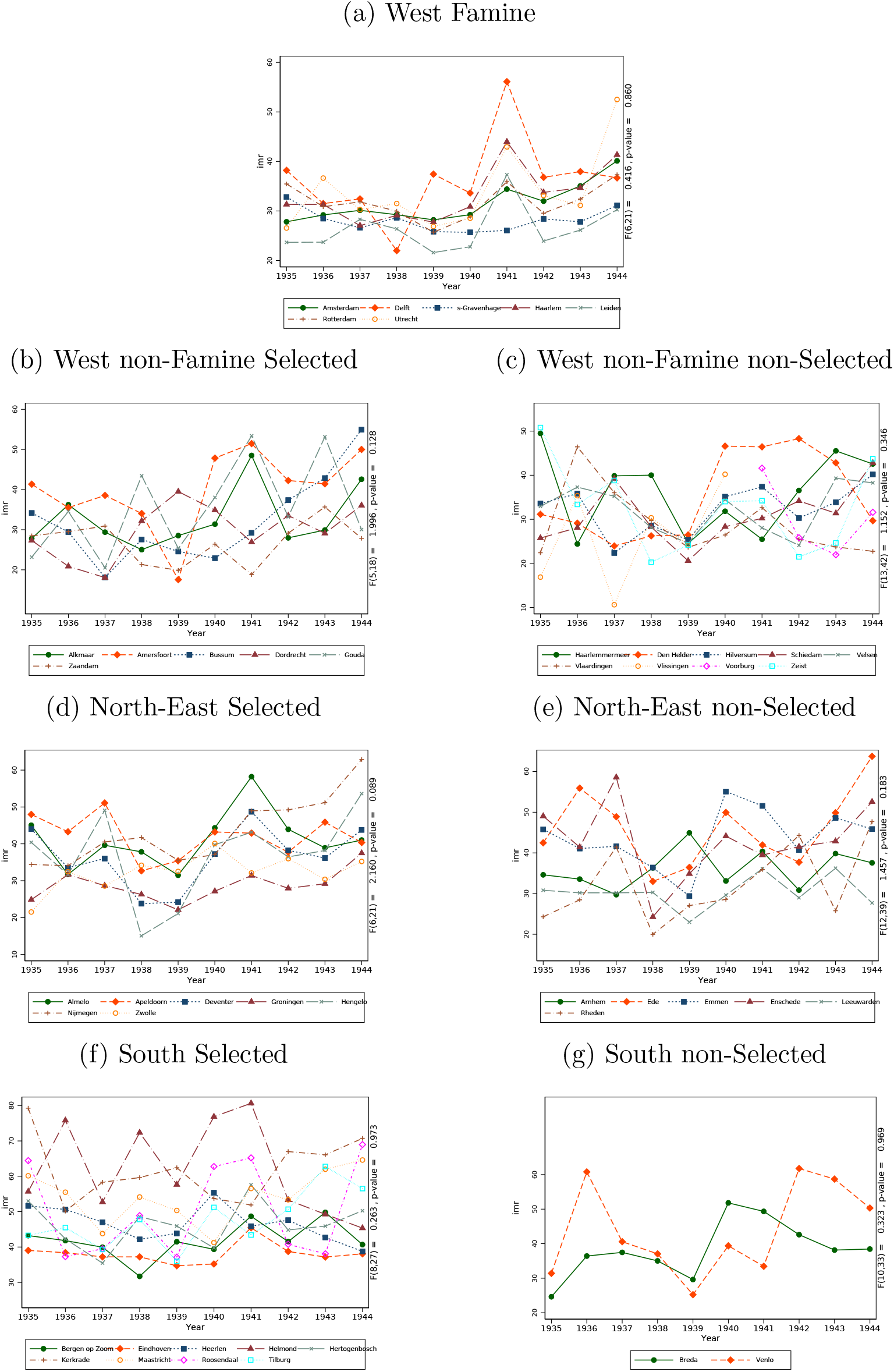
Pretrends for Infant Mortality Rate Note: Pretrends shown for groups of selected, and non-selected, in the analysis cities. A Wald test is performed in each group, by fitting a linear regression with city-specific slopes and testing jointly whether the slopes are equal.

**Figure B6:**
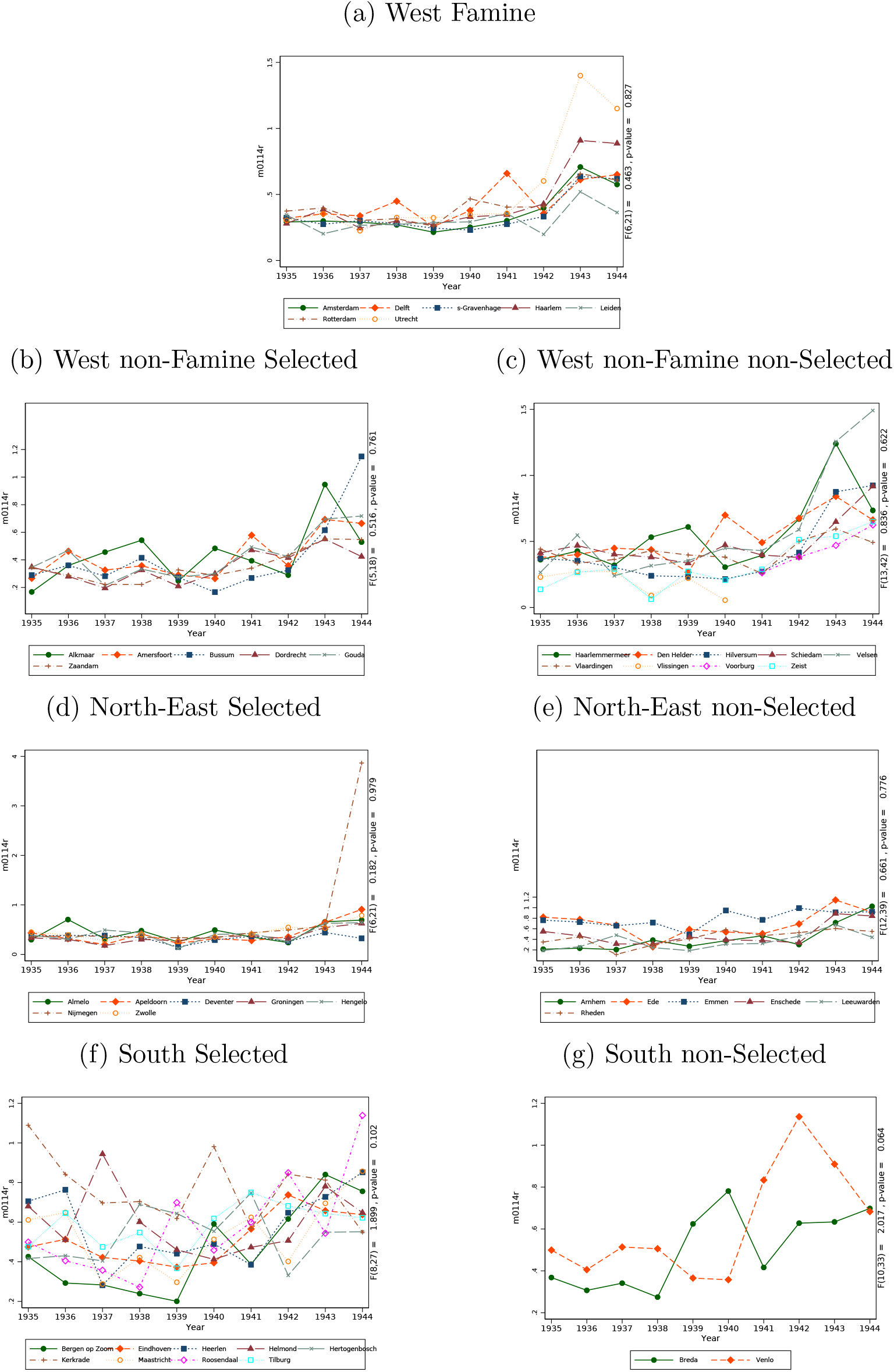
Pretrends for Crude Death Rate: Age 1-14 years Note: Pretrends shown for groups of selected, and non-selected, in the analysis cities. A Wald test is performed in each group, by fitting a linear regression with city-specific slopes and testing jointly whether the slopes are equal.

**Figure B7:**
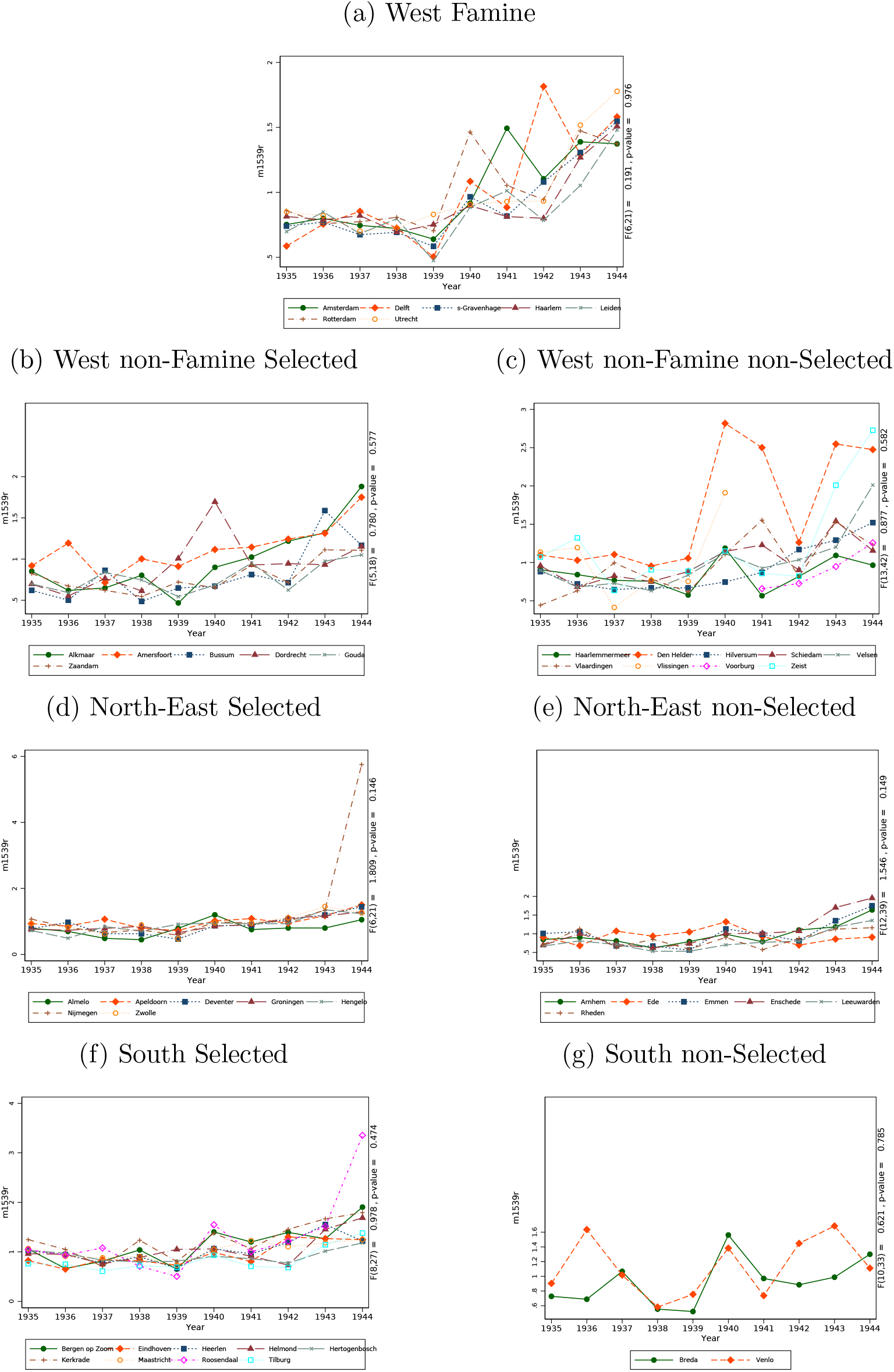
Pretrends for Crude Death Rate: Age 15-39 years Note: Pretrends shown for groups of selected, and non-selected, in the analysis cities. A Wald test is performed in each group, by fitting a linear regression with city-specific slopes and testing jointly whether the slopes are equal.

**Figure B8:**
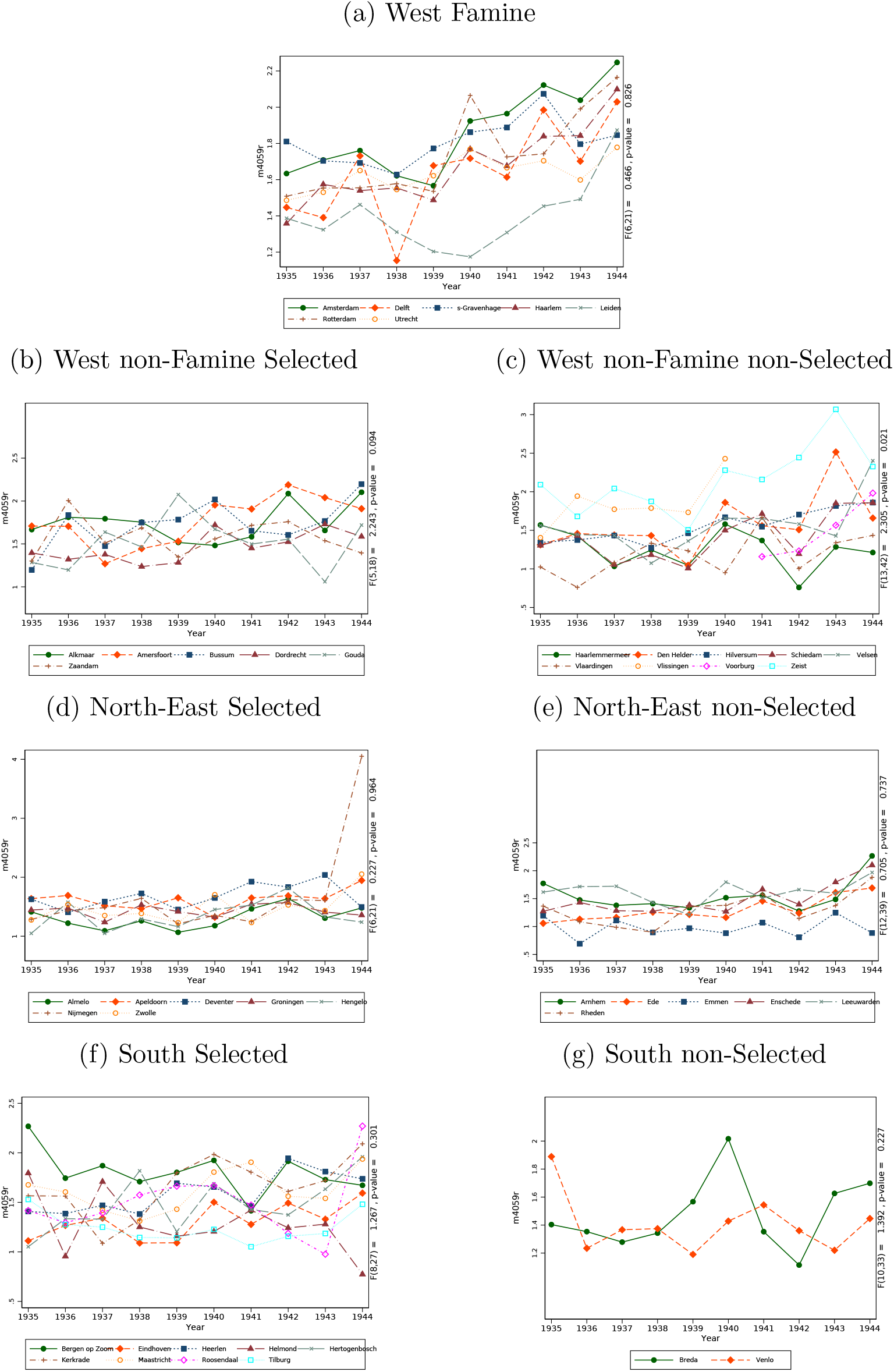
Pretrends for Crude Death Rate: Age 40-59 years Note: Pretrends shown for groups of selected, and non-selected, in the analysis cities. A Wald test is performed in each group, by fitting a linear regression with city-specific slopes and testing jointly whether the slopes are equal.

**Figure B9:**
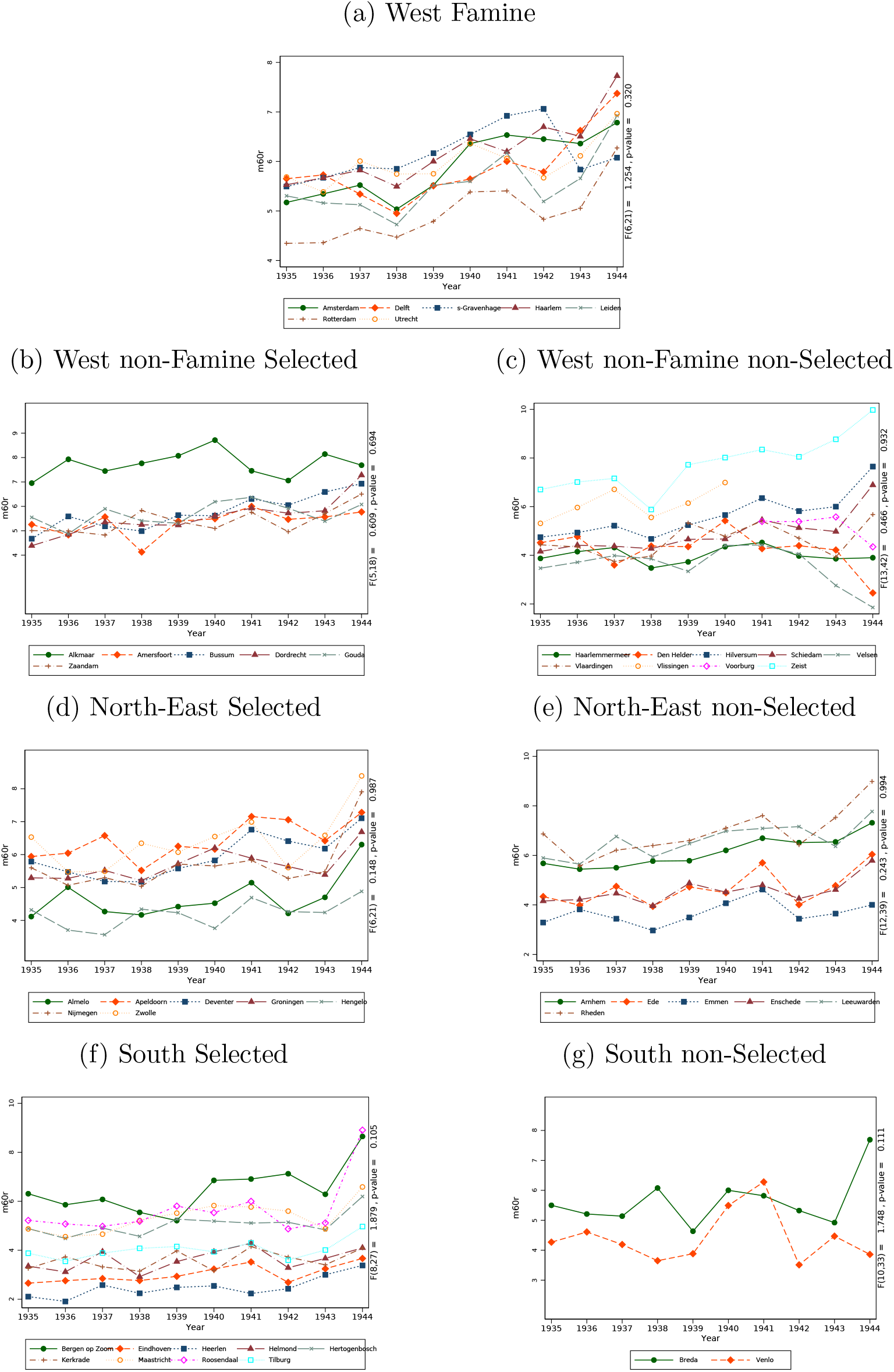
Pretrends for Crude Death Rate: Age 60+ years Note: Pretrends shown for groups of selected, and non-selected, in the analysis cities. A Wald test is performed in each group, by fitting a linear regression with city-specific slopes and testing jointly whether the slopes are equal.

### Appendix C Sensitivity analysis

**Figure C1:**
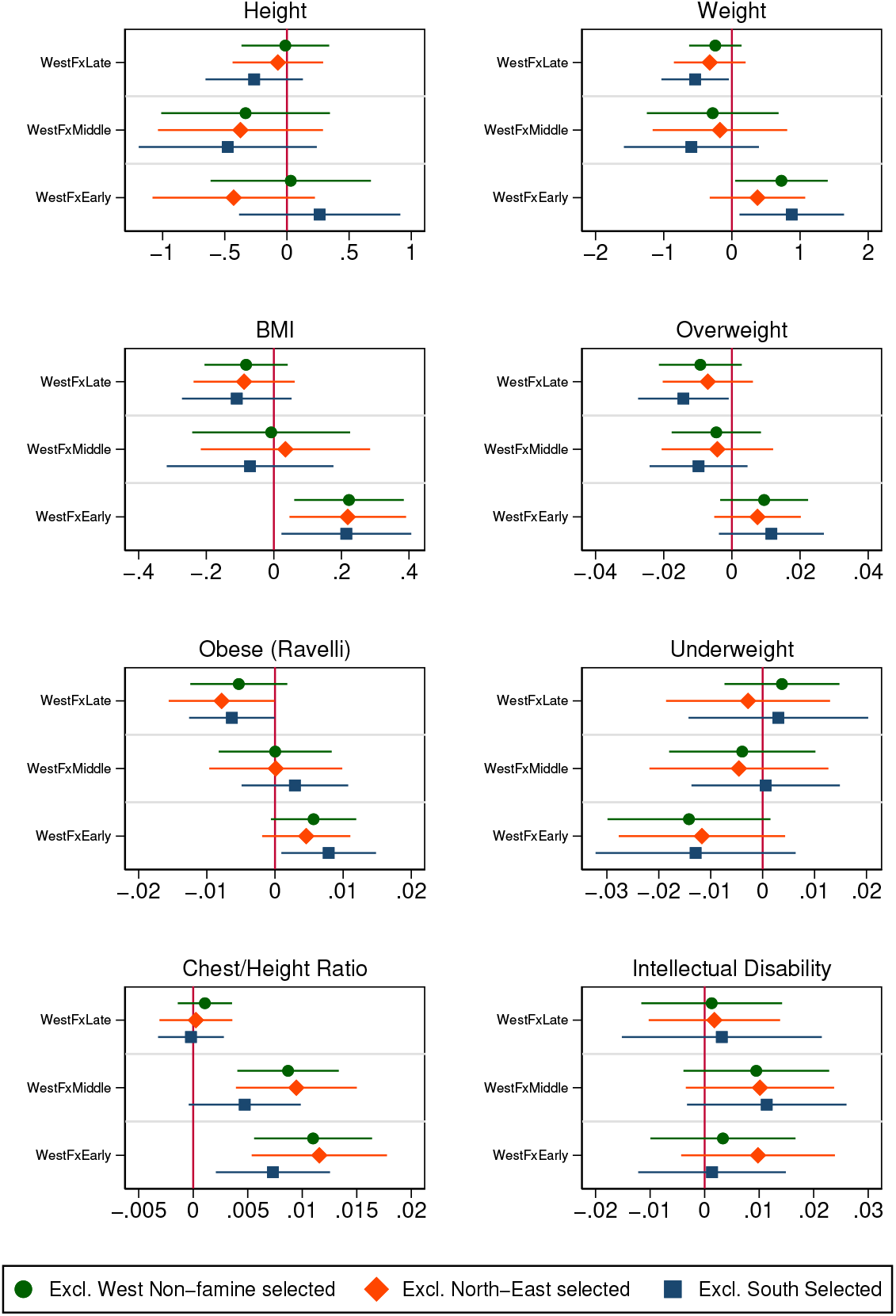
Difference-in-Differences model, excluding one region at a time Note: For each model, the three interaction term estimates are presented along with the 95% Confidence Interval. A vertical line at zero is added to ease examination of significance. Results correspond to models in Table 3.

**Figure C2:**
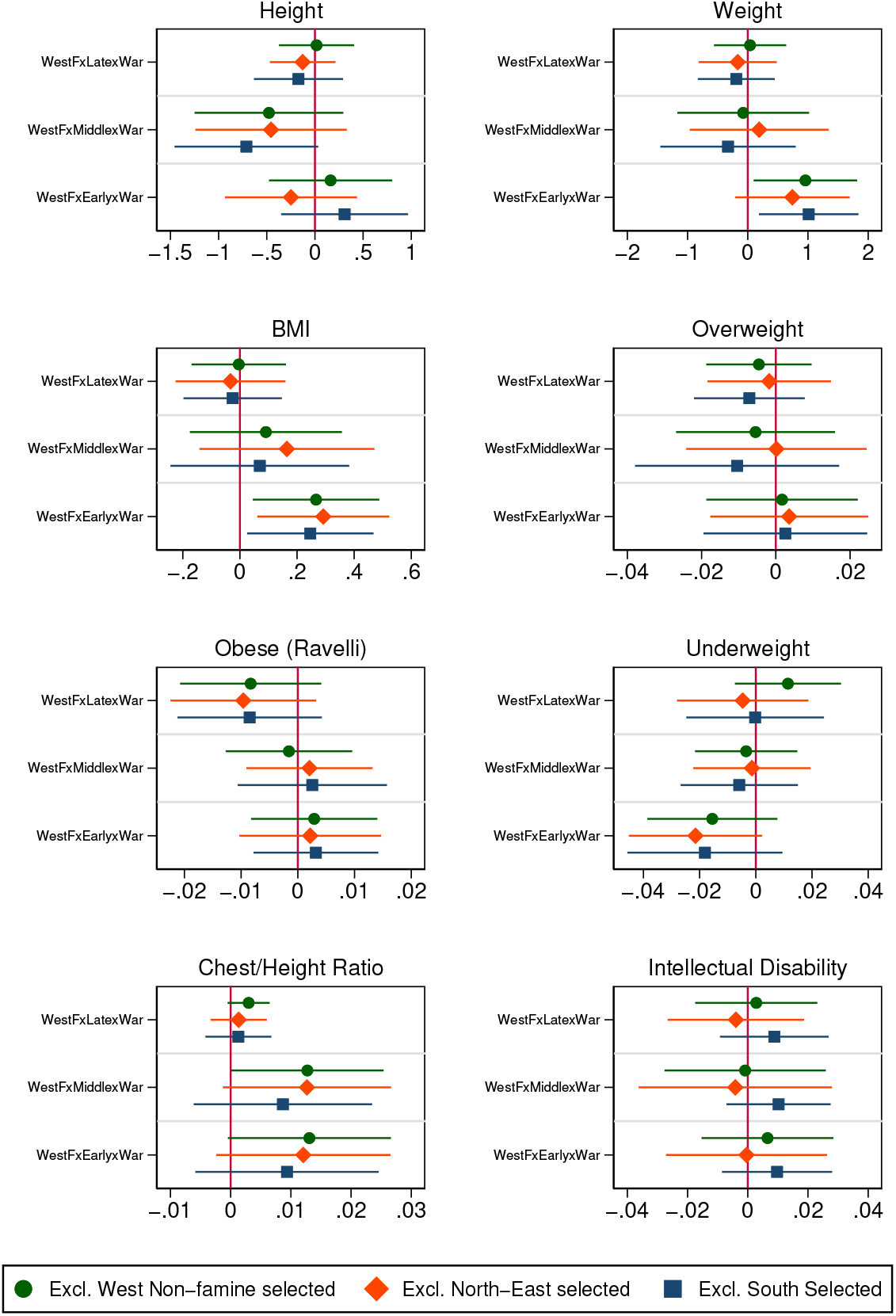
Triple Difference model, excluding one region at a time Note: For each model, the three triple interaction term estimates are presented along with the 95% Confidence Interval. A vertical line at zero is added to ease examination of significance. Results correspond to models in Table 5.

**Figure C3:**
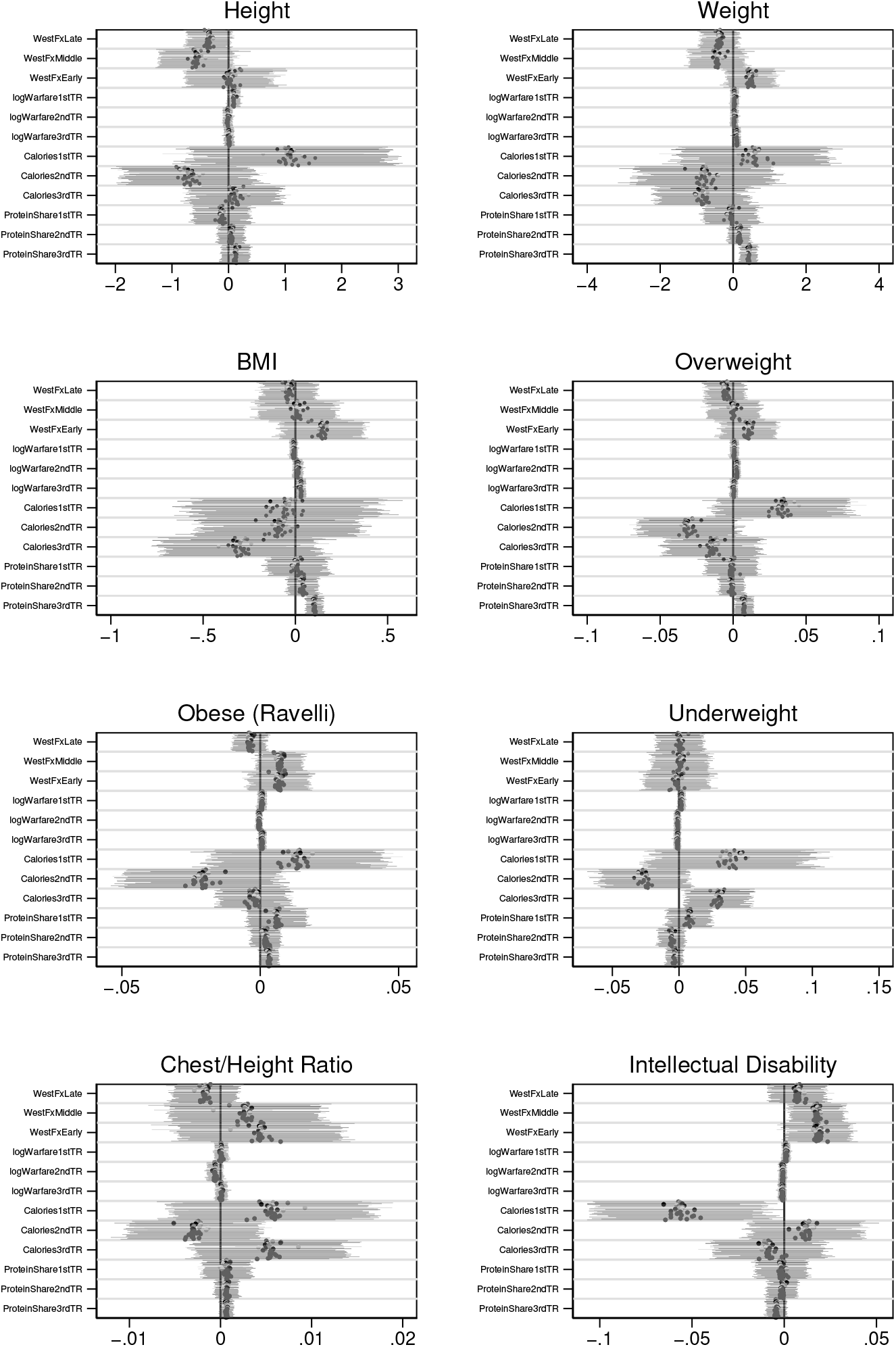
Difference-in-Differences model with mechanisms, excluding one city at a time Note: For each model, all the main estimates are presented along with the 95% Confidence Intervals. A vertical line at zero is added to ease examination of significance. Results correspond to models in Table 6.

### Appendix D Further figures and tables

**Figure D1:**
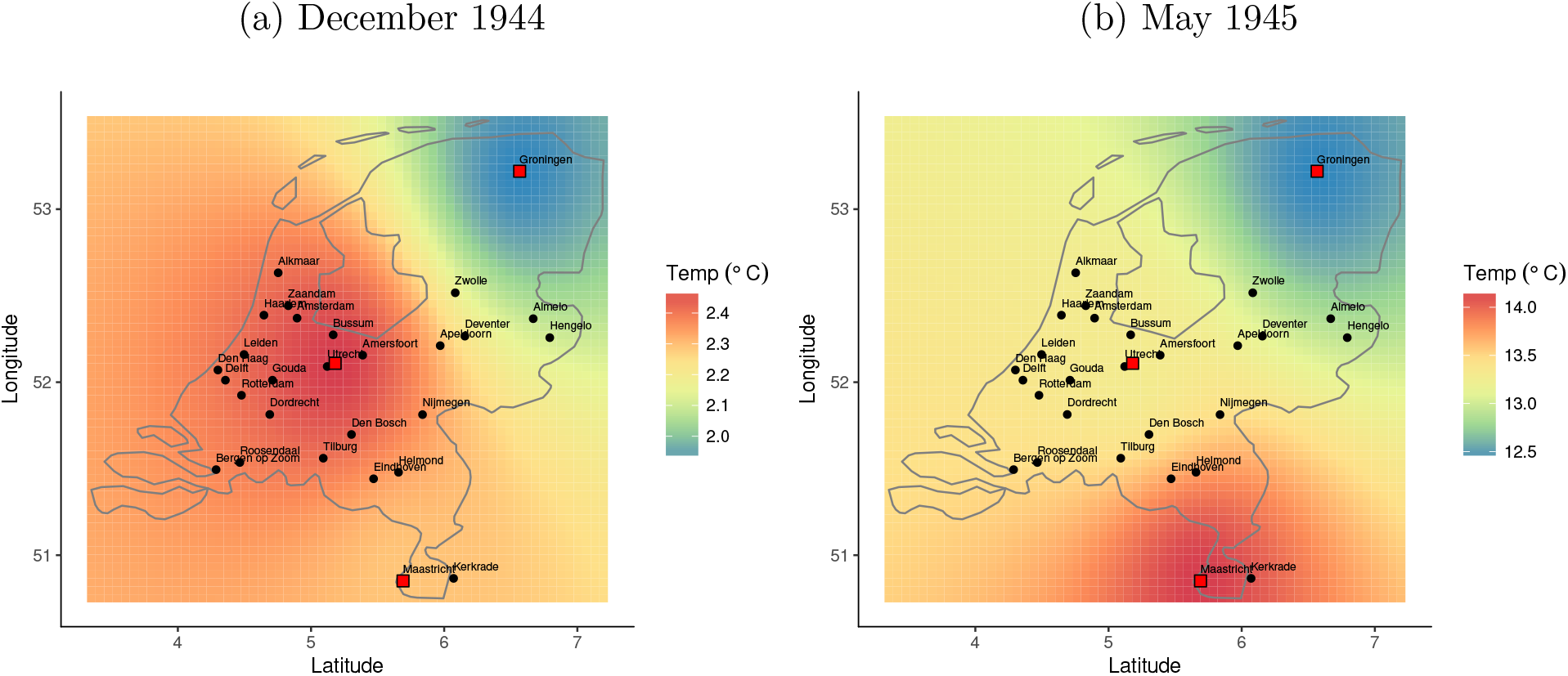
Example of Temperature Interpolation Note: The heatmap shows the predicted temperature across the Netherlands using the Inverse Distance Weighting method. The red squares are the meteorological stations, and the black dots are the cities in the study.

**Figure D2:**
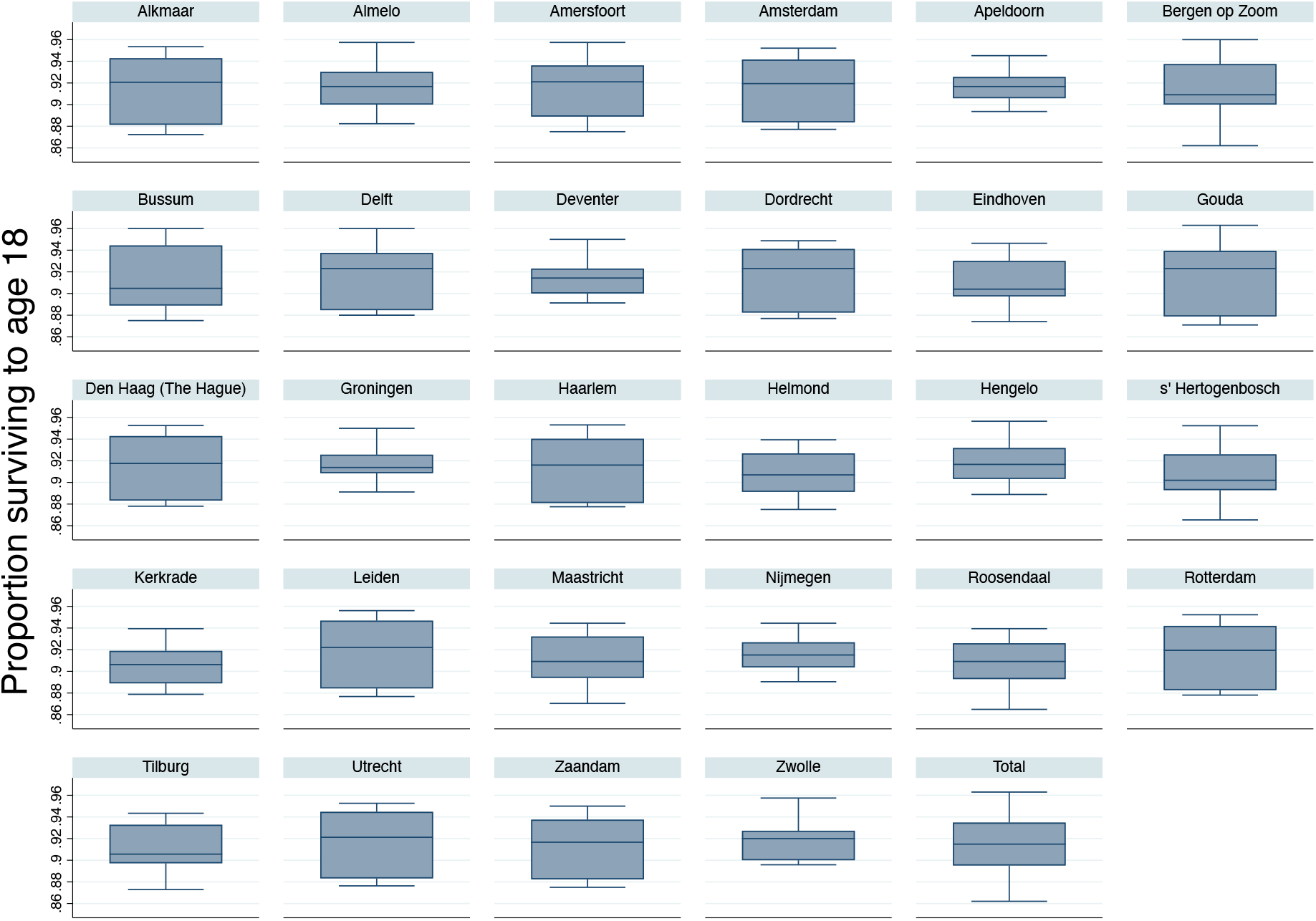
Survival Rates until Age 18 by city Note: Survival rates up to age 18 for those born between May 1944 and July 1945, by city. Source: own calculations based on the recruits data and the information on live births and deaths from Stein et al. (1975) (Table 1).

**Table D1:**
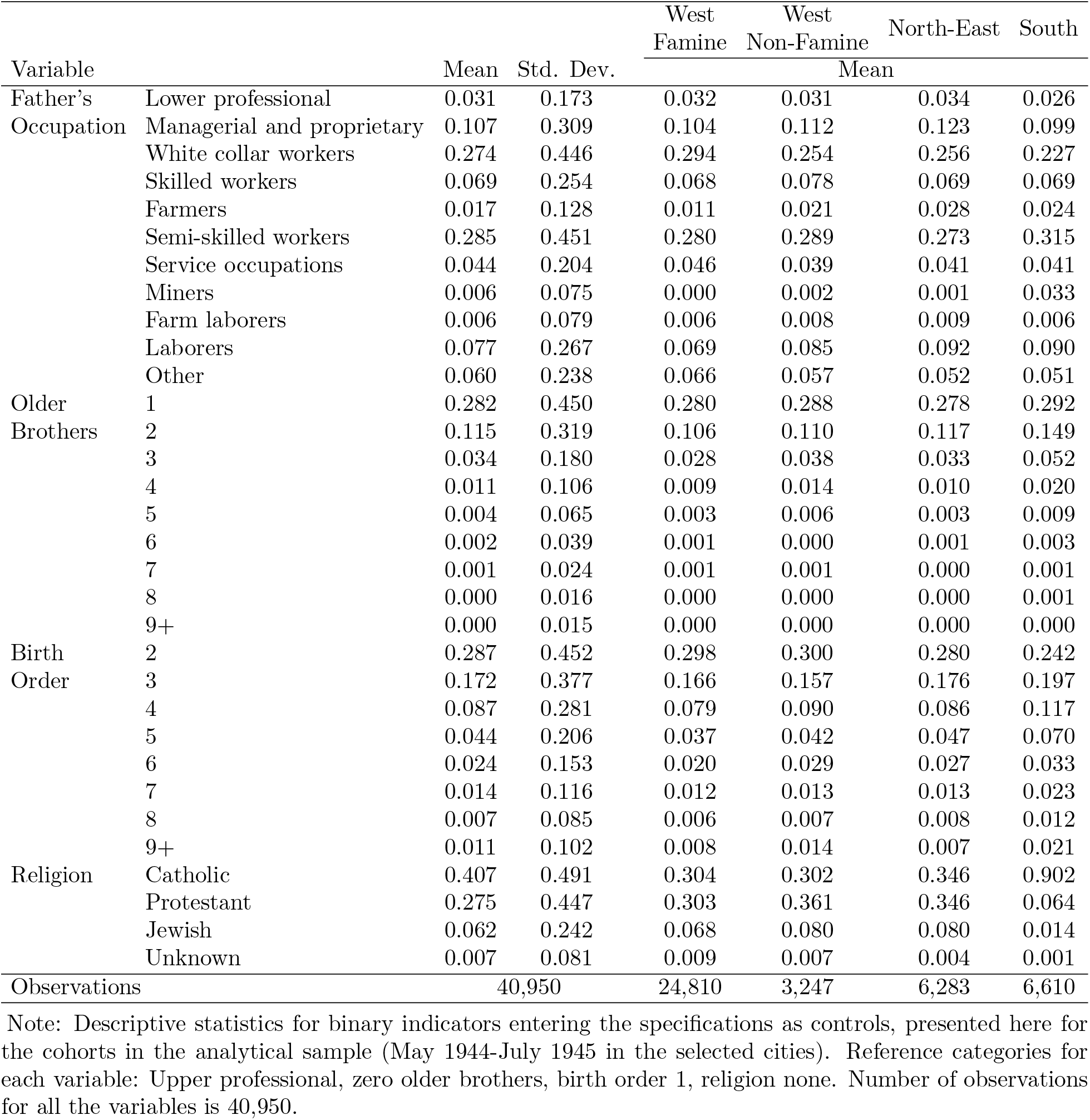
Descriptive Statistics of Control Variables

**Table D2:** Placebo Difference-in-Differences – Ravelli et al. (1976) sample

|  | (1) | (2) | (3) | (4) | (5) | (6) | (7) | (8) |
| --- | --- | --- | --- | --- | --- | --- | --- | --- |
|  | Height | Weight | BMI<br>(Weight/<br>Height<sup>2</sup>) | Overweight<br>(BMI<br>≥25) | Obese<br>(Weight/<br>Height>120%) | Underweight<br>(BMI<<br>18.5) | Chest/<br>Height<br>Ratio | Intellectual<br>Disability<br>(ICD-6 325) |
| WestF×PlaceboLate | -0.132<br>(0.092) | -0.232*<br>(0.134) | -0.040<br>(0.047) | -0.003<br>(0.004) | -0.001<br>(0.003) | -0.002<br>(0.005) | -0.000<br>(0.002) | 0.001<br>(0.005) |
| WestF×PlaceboMiddle | 0.013<br>(0.124) | -0.325**<br>(0.122) | -0.106**<br>(0.040) | -0.003<br>(0.005) | 0.002<br>(0.002) | 0.005<br>(0.005) | -0.003<br>(0.005) | 0.005<br>(0.007) |
| WestF×PlaceboEarly | -0.046<br>(0.113) | -0.251*<br>(0.148) | -0.071*<br>(0.041) | 0.004<br>(0.006) | 0.001<br>(0.002) | 0.007*<br>(0.004) | -0.001<br>(0.005) | -0.001<br>(0.006) |
| Observations | 107,549 | 107,549 | 107,549 | 107,549 | 107,549 | 107,549 | 107,549 | 107,549 |
| Controls and FE | ✓ | ✓ | ✓ | ✓ | ✓ | ✓ | ✓ | ✓ |
| Control Mean | 177.884 | 68.537 | 21.644 | 0.074 | 0.019 | 0.055 | 0.499 | 0.030 |

Note: Robust standard errors in parentheses clustered at the city level. \*\*\* p<0.01, \*\* p<0.05, \* p<0.1. Outcomes, exposures and controls are defined as in Table 4.

**Table D3:**
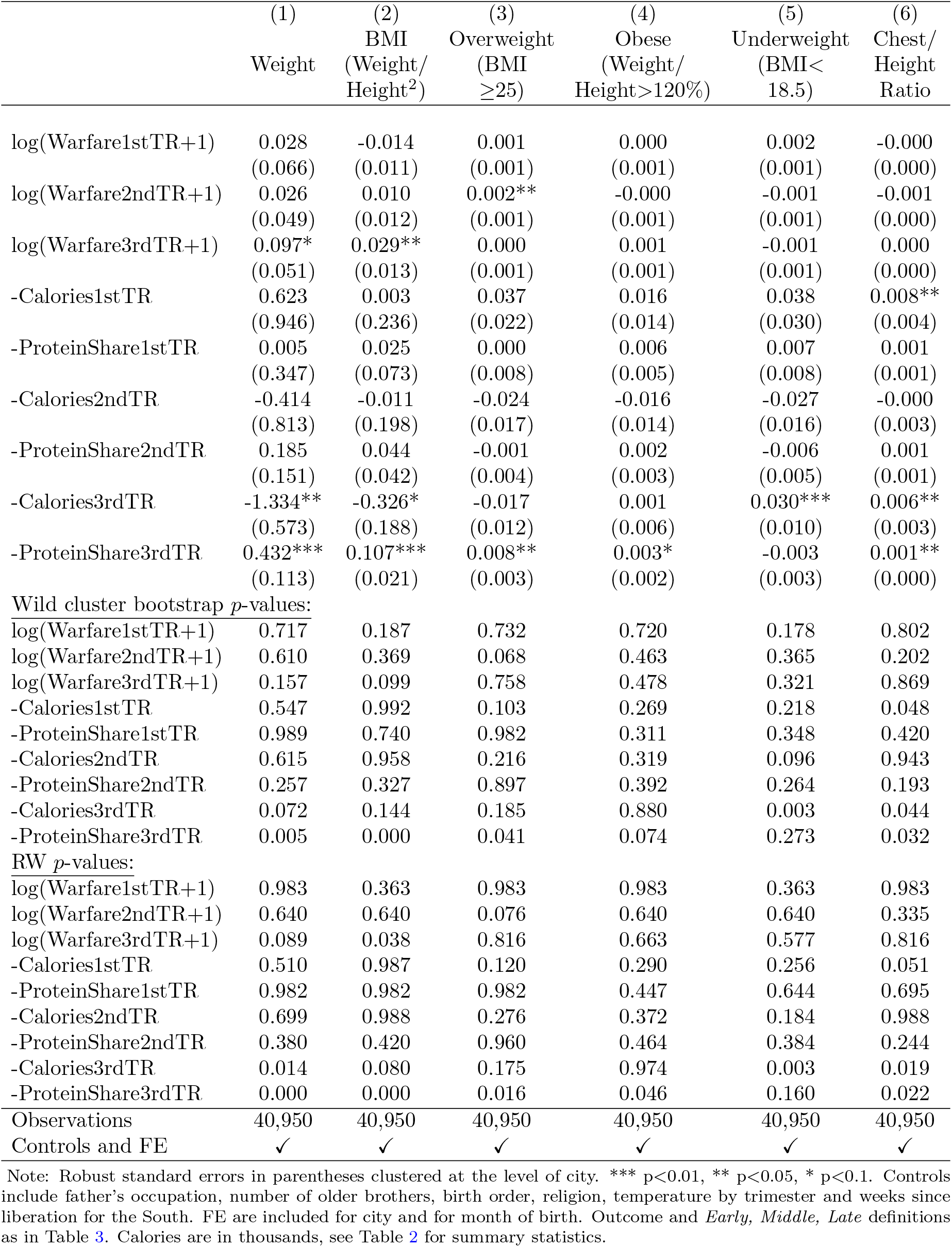
Mechanisms of the Dutch Hunger Winter Effects on Age 18 Outcomes

|  | (1)<br>Weight | (2)<br>BMI<br>(Weight/<br>Height<sup>2</sup>) | (3)<br>Overweight<br>(BMI<br>≥25) | (4)<br>Obese<br>(Weight/<br>Height>120%) | (5)<br>Underweight<br>(BMI<<br>18.5) | (6)<br>Chest/<br>Height<br>Ratio |
| --- | --- | --- | --- | --- | --- | --- |
| log(Warfare1stTR+1) | 0.028<br>(0.066) | -0.014<br>(0.011) | 0.001<br>(0.001) | 0.000<br>(0.001) | 0.002<br>(0.001) | -0.000<br>(0.000) |
| log(Warfare2ndTR+1) | 0.026<br>(0.049) | 0.010<br>(0.012) | 0.002**<br>(0.001) | -0.000<br>(0.001) | -0.001<br>(0.001) | -0.001<br>(0.000) |
| log(Warfare3rdTR+1) | 0.097*<br>(0.051) | 0.029**<br>(0.013) | 0.000<br>(0.001) | 0.001<br>(0.001) | -0.001<br>(0.001) | 0.000<br>(0.000) |
| -Calories1stTR | 0.623<br>(0.946) | 0.003<br>(0.236) | 0.037<br>(0.022) | 0.016<br>(0.014) | 0.038<br>(0.030) | 0.008**<br>(0.004) |
| -ProteinShare1stTR | 0.005<br>(0.347) | 0.025<br>(0.073) | 0.000<br>(0.008) | 0.006<br>(0.005) | 0.007<br>(0.008) | 0.001<br>(0.001) |
| -Calories2ndTR | -0.414<br>(0.813) | -0.011<br>(0.198) | -0.024<br>(0.017) | -0.016<br>(0.014) | -0.027<br>(0.016) | -0.000<br>(0.003) |
| -ProteinShare2ndTR | 0.185<br>(0.151) | 0.044<br>(0.042) | -0.001<br>(0.004) | 0.002<br>(0.003) | -0.006<br>(0.005) | 0.001<br>(0.001) |
| -Calories3rdTR | -1.334**<br>(0.573) | -0.326*<br>(0.188) | -0.017<br>(0.012) | 0.001<br>(0.006) | 0.030***<br>(0.010) | 0.006**<br>(0.003) |
| -ProteinShare3rdTR | 0.432***<br>(0.113) | 0.107***<br>(0.021) | 0.008**<br>(0.003) | 0.003*<br>(0.002) | -0.003<br>(0.003) | 0.001**<br>(0.000) |
| Wild cluster bootstrap *p*-values: |  |  |  |  |  |  |
| log(Warfare1stTR+1) | 0.717 | 0.187 | 0.732 | 0.720 | 0.178 | 0.802 |
| log(Warfare2ndTR+1) | 0.610 | 0.369 | 0.068 | 0.463 | 0.365 | 0.202 |
| log(Warfare3rdTR+1) | 0.157 | 0.099 | 0.758 | 0.478 | 0.321 | 0.869 |
| -Calories1stTR | 0.547 | 0.992 | 0.103 | 0.269 | 0.218 | 0.048 |
| -ProteinShare1stTR | 0.989 | 0.740 | 0.982 | 0.311 | 0.348 | 0.420 |
| -Calories2ndTR | 0.615 | 0.958 | 0.216 | 0.319 | 0.096 | 0.943 |
| -ProteinShare2ndTR | 0.257 | 0.327 | 0.897 | 0.392 | 0.264 | 0.193 |
| -Calories3rdTR | 0.072 | 0.144 | 0.185 | 0.880 | 0.003 | 0.044 |
| -ProteinShare3rdTR | 0.005 | 0.000 | 0.041 | 0.074 | 0.273 | 0.032 |
| RW *p*-values: |  |  |  |  |  |  |
| log(Warfare1stTR+1) | 0.983 | 0.363 | 0.983 | 0.983 | 0.363 | 0.983 |
| log(Warfare2ndTR+1) | 0.640 | 0.640 | 0.076 | 0.640 | 0.640 | 0.335 |
| log(Warfare3rdTR+1) | 0.089 | 0.038 | 0.816 | 0.663 | 0.577 | 0.816 |
| -Calories1stTR | 0.510 | 0.987 | 0.120 | 0.290 | 0.256 | 0.051 |
| -ProteinShare1stTR | 0.982 | 0.982 | 0.982 | 0.447 | 0.644 | 0.695 |
| -Calories2ndTR | 0.699 | 0.988 | 0.276 | 0.372 | 0.184 | 0.988 |
| -ProteinShare2ndTR | 0.380 | 0.420 | 0.960 | 0.464 | 0.384 | 0.244 |
| -Calories3rdTR | 0.014 | 0.080 | 0.175 | 0.974 | 0.003 | 0.019 |
| -ProteinShare3rdTR | 0.000 | 0.000 | 0.016 | 0.046 | 0.160 | 0.022 |
| Observations | 40,950 | 40,950 | 40,950 | 40,950 | 40,950 | 40,950 |
| Controls and FE | ✓ | ✓ | ✓ | ✓ | ✓ | ✓ |

Note: Robust standard errors in parentheses clustered at the level of city. \*\*\* $p < 0.01$ , \*\* $p < 0.05$ , \* $p < 0.1$ . Controls include father's occupation, number of older brothers, birth order, religion, temperature by trimester and weeks since liberation for the South. FE are included for city and for month of birth. Outcome and *Early*, *Middle*, *Late* definitions as in Table 3. Calories are in thousands, see Table 2 for summary statistics.

**Table D4:**
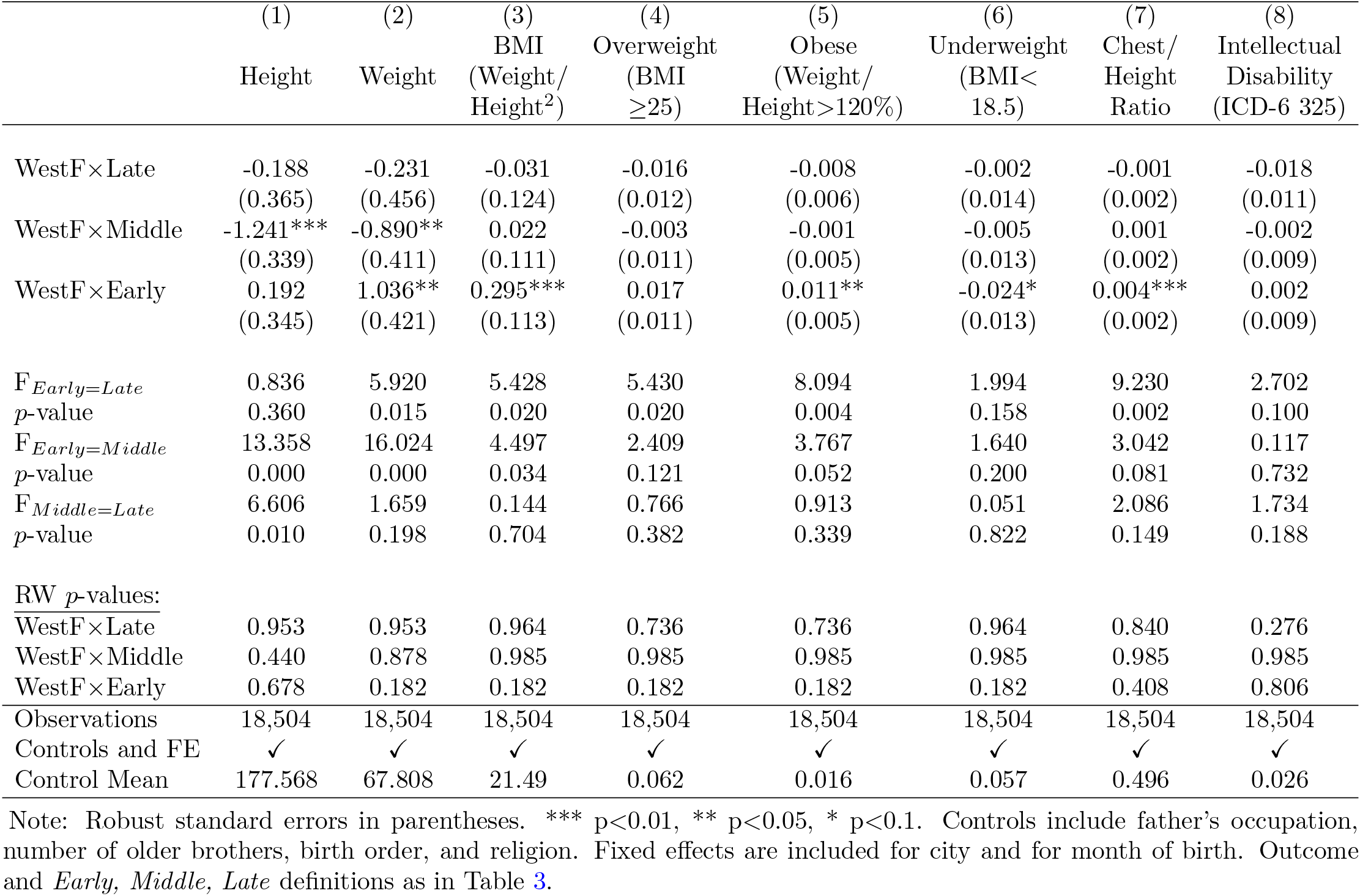
Difference-in-Differences: Age 18 Outcomes, Birth Data cities only

|  | (1) | (2) | (3) | (4) | (5) | (6) | (7) | (8) |
| --- | --- | --- | --- | --- | --- | --- | --- | --- |
|  | Height | Weight | BMI<br>(Weight/<br>Height<sup>2</sup>) | Overweight<br>(BMI<br>≥25) | Obese<br>(Weight/<br>Height>120%) | Underweight<br>(BMI<<br>18.5) | Chest/<br>Height<br>Ratio | Intellectual<br>Disability<br>(ICD-6 325) |
| WestF×Late | -0.188<br>(0.365) | -0.231<br>(0.456) | -0.031<br>(0.124) | -0.016<br>(0.012) | -0.008<br>(0.006) | -0.002<br>(0.014) | -0.001<br>(0.002) | -0.018<br>(0.011) |
| WestF×Middle | -1.241***<br>(0.339) | -0.890**<br>(0.411) | 0.022<br>(0.111) | -0.003<br>(0.011) | -0.001<br>(0.005) | -0.005<br>(0.013) | 0.001<br>(0.002) | -0.002<br>(0.009) |
| WestF×Early | 0.192<br>(0.345) | 1.036**<br>(0.421) | 0.295***<br>(0.113) | 0.017<br>(0.011) | 0.011**<br>(0.005) | -0.024*<br>(0.013) | 0.004***<br>(0.002) | 0.002<br>(0.009) |
| F<sub>Early=Late</sub><br>p-value | 0.836<br>0.360 | 5.920<br>0.015 | 5.428<br>0.020 | 5.430<br>0.020 | 8.094<br>0.004 | 1.994<br>0.158 | 9.230<br>0.002 | 2.702<br>0.100 |
| F<sub>Early=Middle</sub><br>p-value | 13.358<br>0.000 | 16.024<br>0.000 | 4.497<br>0.034 | 2.409<br>0.121 | 3.767<br>0.052 | 1.640<br>0.200 | 3.042<br>0.081 | 0.117<br>0.732 |
| F<sub>Middle=Late</sub><br>p-value | 6.606<br>0.010 | 1.659<br>0.198 | 0.144<br>0.704 | 0.766<br>0.382 | 0.913<br>0.339 | 0.051<br>0.822 | 2.086<br>0.149 | 1.734<br>0.188 |
| RW p-values: |  |  |  |  |  |  |  |  |
| WestF×Late | 0.953 | 0.953 | 0.964 | 0.736 | 0.736 | 0.964 | 0.840 | 0.276 |
| WestF×Middle | 0.440 | 0.878 | 0.985 | 0.985 | 0.985 | 0.985 | 0.985 | 0.985 |
| WestF×Early | 0.678 | 0.182 | 0.182 | 0.182 | 0.182 | 0.182 | 0.408 | 0.806 |
| Observations | 18,504 | 18,504 | 18,504 | 18,504 | 18,504 | 18,504 | 18,504 | 18,504 |
| Controls and FE | ✓ | ✓ | ✓ | ✓ | ✓ | ✓ | ✓ | ✓ |
| Control Mean | 177.568 | 67.808 | 21.49 | 0.062 | 0.016 | 0.057 | 0.496 | 0.026 |

Note: Robust standard errors in parentheses. \*\*\* p<0.01, \*\* p<0.05, \* p<0.1. Controls include father's occupation, number of older brothers, birth order, and religion. Fixed effects are included for city and for month of birth. Outcome and *Early*, *Middle*, *Late* definitions as in Table 3.

**Table D5:**
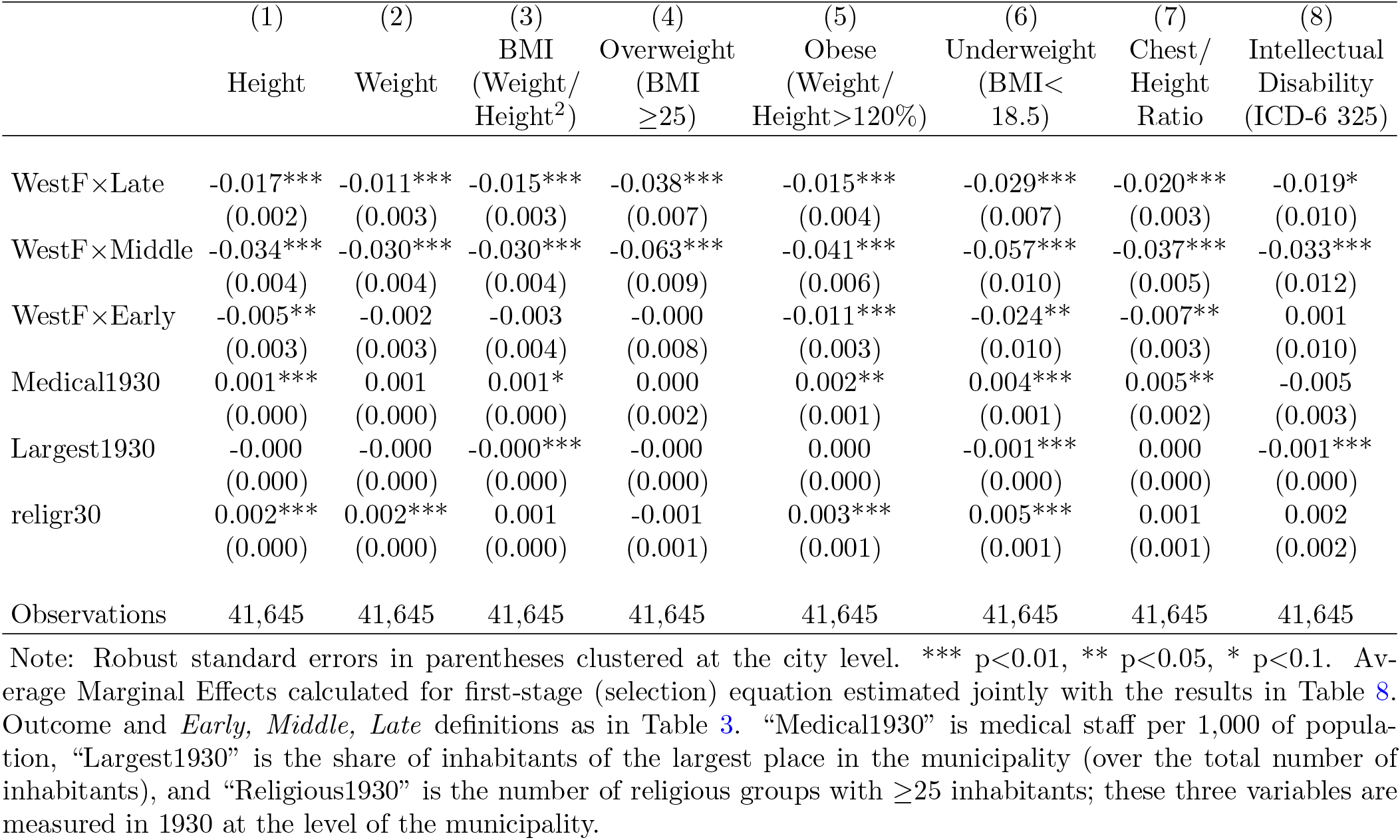
Selection equation Average Marginal Effects of copula selection models

### Appendix E Simulation Study

We conduct the following simulation study to assess the performance of different selection models in a case like ours, with the exposure at aggregate level and the outcomes at individual level. For regions *j* = 1*, …, n_j_* and individuals *i* = 1*, …, n_i_*, we have a sample size of *n_j_ × n_i_* observations clustered in *n_j_* regions. The exposure variable is modelled as *f_j_ ∼ Bernoulli*(0.5). The error terms are modelled to have a Clayton copula structure (see Winkelmann (2012) for the construction method) with dependence parameters *θ_u_* and *θ_ε_*, for the regional-level and individual-level dependence, respectively; depending on whether we impose independence or not, the parameters are taking values of either 2 or 0. The selection equation for survival is constructed as:

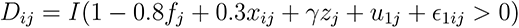

where *f_j_* is a binary variable for famine (1 if famine, 0 otherwise), *x_ij_* is an individual-level exogenous variable, and *z_j_* is a regional-level exogenous variables. We estimate the probability of survival at regional level as 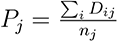 (in all simulations, we keep *n_j_* constant across regions). The individual-level variable (*x_ij_ ∼ N* (0, 2)) is included in both equations, and the region-level variable (*z_j_ ∼ N* (0, 1)) is included only in the selection equation. Finally, the outcome equation is modelled as:

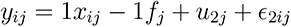

Table E1 shows the results for 100 regions with 100 individuals in each region from six different Data Generating Processes (DGPs), three of them with an exclusion restriction that has a weak relationship with survival (*γ* = 0.05), and three with a strong relationship (*γ* = 0.3). The first panel shows the case with error correlation only at individual level, the middle panel only at regional level, and the bottom panel at both levels. For each scenario, we estimate the outcome equation with OLS before setting the outcome to missing when *D_ij_* = 0 (FULL). We also estimate the outcome equation using GLS estimation (GLS) and correcting for selection using Inverse Probability Weighting (IPW), where the weights are the *P_j_* described above. Finally, we estimate the traditional sample selection model using full maximum likelihood assuming normality in the error terms (HECK) and the sample selection model using the Clayton copula for the dependence of the error terms (COP).

Looking at the results in Table E1 it is evident that the naive OLS estimator is always biased. In the case where the error-term correlation exists only at regional level, the IPW and GLS estimators are unbiased – as expected – but not in the case where there is correlation at the individual level. In fact, the IPW and GLS estimators are performing almost identically across all scenarios. In the case of correlation only at regional level they are also efficient. Finally, in the case of a non-zero correlation in the individual error terms, the best behaved estimators are the HECK and COP. The superiority of COP can be seen in the variance of the estimators, as it is the efficient one. The Standard Deviation is reduced by around 20% across all the scenarios presented in the Table. Moreover, the magnitude of the effect of *z* is affecting the means of both estimators, since both are closer to the true value 1 when *γ* increases.

**Table E1:** Monte Carlo Results, Clayton copula DGP

| Estimator | Weak: $\gamma = 0.05$ | | Strong: $\gamma = 0.3$ | |
| --- | --- | --- | --- | --- |
|  | Mean | Std.Dev. | Mean | Std.Dev. |
| $\sigma_u > 0$ and $\sigma_e = 0$ | | | | |
| FULL | -1.000 | 0.198 | -1.002 | 0.205 |
| OLS | -0.879 | 0.179 | -0.887 | 0.192 |
| IPW | -0.971 | 0.192 | -0.967 | 0.201 |
| GLS | -0.978 | 0.192 | -0.975 | 0.201 |
| HECK | -0.874 | 0.264 | -0.972 | 0.259 |
| COP | -0.922 | 0.192 | -0.942 | 0.205 |
| $\sigma_u = 0$ and $\sigma_e > 0$ | | | | |
| FULL | -0.999 | 0.205 | -0.999 | 0.205 |
| OLS | -0.877 | 0.223 | -0.881 | 0.224 |
| IPW | -0.834 | 0.211 | -0.837 | 0.210 |
| GLS | -0.830 | 0.211 | -0.833 | 0.210 |
| HECK | -0.952 | 0.295 | -0.974 | 0.292 |
| COP | -0.951 | 0.228 | -0.959 | 0.231 |
| $\sigma_u > 0$ and $\sigma_e > 0$ | | | | |
| FULL | -1.000 | 0.199 | -0.998 | 0.202 |
| OLS | -0.754 | 0.164 | -0.761 | 0.172 |
| IPW | -0.802 | 0.166 | -0.803 | 0.173 |
| GLS | -0.808 | 0.166 | -0.809 | 0.173 |
| HECK | -0.909 | 0.287 | -1.010 | 0.244 |
| COP | -0.935 | 0.186 | -0.943 | 0.191 |

Note: Results based on sample size of 10,000 and 2,000 replications.

## Footnotes

1 Famine exposure is not the only design used to study the impact of prenatal malnutrition. For example, the fasting observed during the Ramadan by the Muslim population has been also studied, with mixed results (Almond and Mazumder, 2011; Jürges, 2015; Majid, 2015)

2 Van den Berg and Lindeboom (2018), in their critical review of the literature on famines and health, describe it as the ‘textbook example’.

3 As noted in Van den Berg and Lindeboom (2018), ‘famine often accompany other types of societal disruption. If this point is ignored, this affects the interpretation of the famine as an effect through under-nutrition.

4 Like the majority of the studies in this area, we don’t have information on the nutritional status of the household, hence our estimates are intention-to-treat. However, even in cases where such information is available (e.g. retrospective self-reported hunger episodes like in Van den Berg et al. (2016), exposure to a famine has been used as instrument for nutritional status – a case in which the exclusion restriction might be violated.

5 There is evidence that recall of some adverse childhood events is trustworthy (Havari and Mazzonna, 2015).

6 de Rooij et al. (2010) found that men and women exposed prenatally performed worse on a selective attention task, but this finding was not replicated in another birth cohort with a more comprehensive evaluation of cognitive performance (de Groot et al., 2011).

7 Another famine, also widely studied, occurred in China and lasted for three years between 1959 and 1961 as a result of various economic and social reforms implemented by the government, known as the Great Leap Forward. Li and Lumey (2017) provide a systematic review and meta-analysis of the studies on the long-term health effects of the Chinese famine.

8 See Lumey et al. (2007) for various alternative definitions of treatment and control cohorts in the context of the Dutch Famine.

9 The rationale for doing this is that both rurality and population size are historically major determinants of susceptibility to famine. For example, Breda, Leeuwarden, Venlo and Vlaardingen underwent municipal merges (in 1942, 1944, 1941 and 1942, respectively), which increased their population size; den Helder, Velsen and Vlissingen were evacuated in 1941, 1942 and 1940 respectively, because the use of their harbours as German naval bases implied a heightened risk of the bombing by the allied for the civilians.

10 The estimated regression is *y* = *β*_0_ + *β*_1_*year* + *β*_2_*_,k_city_k_* + *β*_3_*_,k_city_k_ × year* where *y* is the corresponding outcome, *year* takes values from 1935 to 1939, and *city_k_* is a dummy variables for each city *k* = 1*, …, K* for the *K* cities included in each group. Then, the Wald test is performed on whether all *β*_3_*_,k_* are equal to 0.

11 The shares were calculated using the following standard formulae: *Protein share* = (*Protein* (*grams*) *×* 4)*/Calories* (*kcals*), *Fat share* = 9 *× Fat*(*grams*)*/Calories*(*kcals*), and (*Carbohydrates share* = 100 *−*(*Protein share* + *Fat share*)).

12 The winter of 1944-45 was assigned a Hellman cold index of 83.3 by the Dutch national meteorological institute (KNMI), which puts it at the 37th position in the ranking of coldest winters in the Netherlands since 1901. There was a period (from January 23-30, 1945) with seven ice days (maximum temperature below 0°C) and a lowest temperature of −13.3°C (Ekamper et al., 2017).

13 As noted in Van den Berg and Lindeboom (2018), ‘one of the reasons why the Dutch famine has been widely used in the literature is that it was so short that the quality of the population registers for the exposed cohorts was not heavily affected. Indeed, we can rule out potential measurement error in the date of birth: when we plot the distribution of the calendar day of birth for all the recruits in the years 1944-1947, we see no sign of heaping, both overall, by month, and by city (graphs available upon request).

14 In the analysis, we transform both calories and protein shares to their negative values, so that their coefficients can be interpreted as the effects of a one-unit decrease. Additionally, we convert the calories in thousands to ease readability of the estimated coefficients; and we use a log(x+1) transformation for the warfare variables, due to the high skewness and to avoid loosing the observations with zero deaths.

15 Unfortunately, we don’t have information on miscarriages. However, in Table 7 we show that there is no effect of in utero famine exposure on sex ratios: and given that males are more vulnerable to maternal stress in pregnancy, we can rule out famine impacts on fetal deaths (see Sanders and Stoecker (2015) for a related use of sex ratio of live births as indicator of fetal deaths).

16 For a subset in two cities (Amsterdam and Tilburg) it was also demonstrated that deaths and out-migrations were low and no births were missing from the military register.

17 We have data from one hospital per city, with the exception of Amsterdam for which we have data for two hospitals.

18 Table D1 in the Appendix presents descriptive statistics of these variables. In our analytical sample, 28% of the cohort members had fathers who were semi-skilled workers, whereas 27% had fathers who were white collar workers. 35% were first-born, 29% second-born and 17% third-born, with 45% having at least one older brother. Catholics constituted 40% of the sample and Protestants 28%.

19 Note that we have the entire population of male births in the study period. Outcome differences between subpopulations defined by some attributes should simply be estimates which would be known with certainty (i.e., the standard errors should be zero). By reporting statistical significance nevertheless, we implicitly assume that there is a superpopulation from which the population is randomly sampled. As with samples drawn from the population, uncertainty in our case emerges from the unobservability of the superpopulation – this may be, for example, future populations, in which the uncertainty would emerge from year-to-year variation (for most recent discussion on the issue see Abadie et al. (2020)).

20 The results reported here are based on 5,000 replications for the wild cluster bootstrap and on 1,000 replications for the Romano and Wolf procedure; the results are not sensitive to different numbers of replications.

21 Mobility is unlikely to be an issue in our setup. First, as mentioned in 3.1, we exclude from our control group the cities which have been evacuated; second, as mentioned in 3.2, Stein et al. (1975) showed that out-migration was not an issue.

22 While a general implementation can be traced back to Lee (1983), the use of copulas in this context has become explicit since Smith (2003).

23 Note that here we define obesity as weight/height*>*120, as used by Ravelli, and not with the current definition of BMI *≥*30.

24 The only exception is the coefficient on the chest-height ratio for mid-pregnancy exposure, which is only significant at 10% level.

25 Calories and protein shares have been transformed into negative values, so the effect can be interpreted as reductions in each. Temperature and weeks since liberation in the South are also controlled for in every specification (they are not shown in the table since their associated coefficients fail to achieve statistical significance). Table D3 in the Appendix presents the same specification without the Difference-in-Differences interaction terms.

26 Instead, the famine effects on underweight are driven to zero once we account for the reduction in calories in the third trimester, highlighting how changes in different parts of the BMI distribution might be driven by different determinants.

27 For example, in some families pregnant women might have been protected, by allocating them a higher share of resources than the one implied by the rations.

28 The zero effect on sex ratio for the Dutch cohorts is shown and discussed in Cramer and Lumey (2010). Other studies, instead, have found significant impacts of early shocks on sex ratios, see for example Catalano et al. (2006), Almond et al. (2007) and Sanders and Stoecker (2015).

29 See Winkelmann (2012) and Gomes et al. (2018) for simulation studies on the selection of copula structures.

30 The results for the selection equation are presented in Table D5. The exclusion restrictions have the expected sign: cities with more medical staff per capita and more religious groups in 1930 have higher survival rates, while those with a denser population structure have lower ones, both for reduced availability of food and also for easier spread of infections.

31 Given that the dependence parameter does not have the same interpretation across different copulas, we have transformed it into a standard rank correlation coefficient, Kendal’s *τ*.

32 Gørgens et al. (2012) also found that taller children were more likely to survive the Chinese famine.

## Notes

This work was supported by the NIA/NIH grant “Prenatal Under Nutrition and Mortality Through Age 63” (R01AG028593-01A2, PI:LHL) where GC, PE, GB are co-Is. GC and SP have received funding from European Union’s HORIZON 2020 research and innovation programme (grant no. 633595 DynaHEALTH). GC has also received funding from the European Research Council (ERC) under the European Union’s Horizon 2020 research and innovation programme grant agreement No 819752 - DEVORHBIOSHIP - ERC-2018COG.

### Competing Interest Statement

The authors have declared no competing interest.

### Author Declarations

Columbia University IRB gave ethical approval for this work.

